# Mitochondrial genome copy number measured by DNA sequencing in human blood is strongly associated with metabolic traits via cell-type composition differences

**DOI:** 10.1101/2020.10.23.20218586

**Authors:** Liron Ganel, Lei Chen, Ryan Christ, Jagadish Vangipurapu, Erica Young, Indraniel Das, Krishna Kanchi, David Larson, Allison Regier, Haley Abel, Chul Joo Kang, Alexandra Scott, Aki Havulinna, Charleston W. K. Chiang, Susan Service, Nelson Freimer, Aarno Palotie, Samuli Ripatti, Johanna Kuusisto, Michael Boehnke, Markku Laakso, Adam Locke, Nathan O. Stitziel, Ira M. Hall

## Abstract

Mitochondrial genome copy number (MT-CN) varies among humans and across tissues and is highly heritable, but its causes and consequences are not well understood. When measured by bulk DNA sequencing in blood, MT-CN may reflect a combination of the number of mitochondria per cell and cell type composition. Here, we studied MT-CN variation in blood-derived DNA from 19,184 Finnish individuals using a combination of genome (N = 4,163) and exome sequencing (N = 19,034) data as well as imputed genotypes (N = 17,718). We identified two loci significantly associated with MT-CN variation: a common variant at the *MYB-HBS1L* locus (P = 1.6×10^−8^), which has previously been associated with numerous hematological parameters; and a burden of rare variants in the *TMBIM1* gene (P = 3.0×10^−8^), which has been reported to protect against non-alcoholic fatty liver disease. We also found that MT-CN is strongly associated with insulin levels (P = 2.0×10^−21^) and other metabolic syndrome (metS) related traits. Using a Mendelian randomization framework, we show evidence that MT-CN measured in blood is causally related to insulin levels. We then applied an MT-CN polygenic risk score (PRS) derived from Finnish data to the UK Biobank, where the association between the PRS and metS traits was replicated. Adjusting for cell counts largely eliminated these signals, suggesting that MT-CN affects metS via cell type composition. These results suggest that measurements of MT-CN in blood-derived DNA partially reflect differences in cell-type composition and that these differences are causally linked to insulin and related traits.

## Introduction

There are many reported links between mitochondrial content and cardiometabolic phenotypes in various tissues, including adipose^1–3^, liver^1,4,5^, skeletal muscle^1,6–10^, and blood^11–17^. Traits associated with mitochondrial (MT) content include coronary heart disease (CHD), type 2 diabetes, and metabolic syndrome traits such as insulin sensitivity/resistance, obesity, and blood triglycerides. However, these studies have generally been limited by small sample sizes and low statistical power. This, in addition to the use of heterogeneous mitochondrial quantification methods^18^, has led to inconsistencies in the literature about the strength and directions of effect between mitochondrial content and metabolic syndrome (metS) traits. In one large WGS study of mitochondrial genome copy number (MT-CN) in 2,077 Sardinians, Ding *et al*. estimated the heritability of MT-CN at 54% and detected significant associations between MT-CN and both waist circumference and waist-hip ratio, but found no association with body mass index (BMI)^11^. Another large study (N = 5,150) found virtually no evidence of association between qPCR-measured MT-CN and any of several cardiometabolic phenotypes^19^. The only exception was an inverse association with insulin that was identified in one cohort but did not survive meta-analysis across cohorts. However, a study of 21,870 individuals from 3 cohorts showed a significant inverse relationship between MT-CN (measured by microarray probe intensities in two cohorts and qPCR in the third) and incident cardiovascular disease^20^.

Although variations in MT-CN measured from whole blood can in principle be attributed to either variability of MT copy number within cells or the cell type composition of the blood (given that different cell types have varying MT content^21–23^), the literature on this subject is inconclusive. Using CpG methylation data, a large (N = 11,443), low-coverage (1.7x autosomal; 102x mitochondrial) sequencing study of the link between MT-CN and major depressive disorder using buccal DNA from Chinese women concluded that variability of MT-CN from buccal swabs was not due to differences in cell type composition^24^. However, this study did not do a similar experiment in blood. Two small (N = 756 and N = 400) studies identified an association between MT content and CHD that they attributed to variable MT-CN within leukocytes, but they did not directly investigate the possibility of cell type composition being the true driver of the association^12,17^. For brevity, we will use the term “MT-CN” to refer to the underlying phenotype reflected by measuring this quantity for the remainder of this work, with these caveats.

While several studies have found that peripheral blood MT content is heritable, only a small number of MT-CN associated loci have been identified^25–27^. In one of these studies, Curran *et al*. used linkage analysis in Mexican Americans to find an MT-CN associated locus near a marker previously associated with triglyceride levels^26,28,29^, providing further indirect evidence for the link between MT-CN and metabolic syndrome.

Here, we take advantage of large-scale genome, exome, and array genotype data to investigate the causes and effects of MT-CN in a large, deeply phenotyped Finnish cohort. Our results reveal novel links with metabolic syndrome and provide evidence supporting a causal role for MT-CN.

## Material and Methods

### Genotype and phenotype data

Whole genome sequencing (WGS) was performed on a cohort of 4,163 samples comprising 3,074 male samples from the METSIM study^30^ and 1,089 male and female samples from the FINRISK study^31^. Genomic DNA was fragmented on the Covaris LE220 instrument targeting 375 bp inserts. Automated Illumina libraries were constructed with the TruSeq PCR-free (Illumina) or KAPA Hyper PCR-free library prep kit (KAPA Biosystems/Roche) on the SciClone NGS platform (Perkin Elmer). The fragmented genomic DNA was size-selected on the SciClone instrument with AMPure XP beads to tighten the distribution of fragmented DNA to ensure the average insert of the libraries was 350-375 bp. We followed the manufacturer’s protocol as provided by Perkin Elmer, with the following exception: post ligation, the libraries were purified twice with a 0.7x AMPure bead/sample ratio to eliminate any residual adaptors present. An aliquot of the final libraries was diluted 1:20 and quantitated on the Caliper GX instrument (Perkin Elmer). The concentration of each library was accurately determined through qPCR utilizing the KAPA library Quantification Kit according to the manufacturer’s protocol (KAPA Biosystems/Roche) to produce cluster counts appropriate for the Illumina HiSeqX instrument. Libraries were pooled and run over a few lanes of the HiSeq X to ensure the libraries within the pool were equally balanced. The final pool of balanced libraries was loaded over the remaining number of HiSeq X lanes to achieve the desired coverage for this project. 2×150 paired end sequence data were demultiplexed using a single index, which was a restriction on the HiSeqX instrument at this time. A minimum of 19.5x coverage was achieved per sample.

The quality of the aligned sequence data was assessed using metrics generated by Picard^32^ v2.4.1, Samtools^33^ v1.3.1 and VerifyBamID^34^ v1.1.3. Based on the output files from Picard, the following alignment statistics were collected for review: PF_MISMATCH_RATE, PF_READS, PF_ALIGNED_BASES, PCT_ADAPTER, PCT_CHIMERAS, PCT_PF_READS_ALIGNED, PCT_READS_ALIGNED_IN_PAIRS, PF_HQ_ALIGNED_BASES, PF_HQ_ALIGNED_Q20_BASES, PF_HQ_ALIGNED_READS, MEAN_INSERT_SIZE, STANDARD_DEVIATION, MEDIAN_INSERT_SIZE, TOTAL_READS, PCT_10x, and PCT_20x. Alignment rate was calculated as PF_READS_ALIGNED/TOTAL_READS. The formula for haploid coverage was as follows: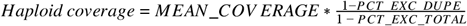 From the Samtools output, inter-chromosomal rate was calculated as: 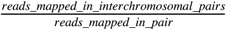 and discordant rate was calculated as: *reads*_*mapped*_*percentage − reads*_*mapped*_*in*_*proper*_*pairs*_*percentage*.

Properly paired percentage (reads_mapped_in_proper_pairs_percentage) and singleton percentage (reads_mapped_as_singleton_percentage) were also reviewed. From VerifyBamID, the Freemix value was reviewed.

The metrics for judgement of passing data quality were: FIRST_OF_PAIR_MISMATCH_RATE < .05, SECOND_OF_PAIR_MISMATCH_RATE < 0.05, haploid coverage ≥ 19.5, interchromosomal rate < .05, and discordant rate < 5. All of the above metrics must have been met in order for the sample to be assigned as QC pass. If a sample did not meet the passing criteria, the following failure analysis was performed: a) If the Freemix score was at least 0.05, the sample or the library was considered contaminated, and both the library and the sample were abandoned; b) if the discordant rate was over 5 and/or the inter-chromosomal rate was over 0.05, the quality of DNA was considered poor and the sample was removed from the sequencing pipeline; and c) in the case of a) and b), the collaborator was contacted to determine if selection of a replacement sample from the same cohort was desired or feasible.

Separately, whole exome sequencing (WES) data (N = 19,034), genotyping array data (N = 17,718) imputed using the Haplotype Reference Consortium panel^35^ v1.1, and transformed, normalized quantitative cardiometabolic trait data were obtained from an earlier study^36^. FINRISK array data came in nine genotyping batches, two of which were excluded from the present study due to small sample size. The traits, normalization and transformation procedures, and sample sizes are described in a previous publication^36^. The WES and imputed sample sets contained 4,013 and 3,929 of the 4,163 WGS samples included in the present study.

All participants in both the METSIM and FINRISK studies provided informed consent, and study protocols were approved by the Ethics Committees at participating institutions (National Public Health Institute of Finland; Hospital District of Helsinki and Uusimaa; Hospital District of Northern Savo). All relevant ethics committees approved this study.

### WGS callset generation and quality control

Single nucleotide polymorphisms (SNPs) and small insertions and deletions were called from the full set of 4,163 samples using GATK^37^ v3.5. GVCFs containing SNVs and Indels from GATK HaplotypeCaller (`-ERC GVCF -GQB 5 -GQB 20 -GQB 60-variant_index_type LINEAR -variant_index_parameter 128000`) were first processed to ensure no GVCF blocks crossed boundaries every 1 Mb (CombineGVCFs; `--breakBandsAtMultiplesOf 1000000`). The resulting GVCFs were then processed in 10 Mb shards across each chromosome. Each shard was combined (CombineGVCFs), genotyped (GenotypeGVCFs; `-stand_call_conf 30-stand_emit_conf 0`), hard filtered to remove alternate alleles uncalled in any individual removed (SelectVariants; `--removeUnusedAlternates`), and hard filtered to remove lines solely reporting symbolic deletions in parallel. All shards were jointly recalibrated (VariantRecalibrator) and then individually filtered (ApplyRecalibration) based on the recalibration results. All of the above methods were performed using GATK v3.5. SNP variant recalibration was performed using the following options to VariantRecalibrator and all resources were drawn from the GATK hg38 resource bundle (v0):

~~~
-mode SNP
-resource:hapmap,known=false,training=true,truth=true,prior=15.0
-resource:omni,known=false,training=true,truth=true,prior=12.0
-resource:1000G,known=false,training=true,truth=false,prior=10.0
-resource:dbsnp,known=true,training=false,truth=false,prior=2.0
-an QD -an DP -an FS -an MQRankSum -an ReadPosRankSum
-tranche 100.0 -tranche 99.9 -tranche 99.0 -tranche 90.0
~~~

Indel variant recalibration was performed using the following options to VariantRecalibrator (with the same resource bundle as with SNPs):

~~~
-mode INDEL
-resource:mills,known=true,training=true,truth=true,prior=12.0
-an DP -an FS -an MQRankSum -an ReadPosRankSum
--maxGaussians 4
-tranche 100.0 -tranche 99.9 -tranche 99.0 -tranche 90.0
~~~

When applying the variant recalibration the following options were used:

For SNPs: `--ts_filter_level 99.0`

For Indels: `--ts_filter_level 99.0`

Following SNP and INDEL variant recalibration, multiallelic variants were decomposed and normalized with vt^38^ v0.5. Duplicate variants and variants with symbolic alleles were subsequently removed. The bottom tranche of variants identified by GATK’s Variant Quality Score Recalibration tool and variants with missingness greater than 2% were removed as well, although variants with allele balance between 0.3 and 0.7 were rescued. Variants with Hardy-Weinberg equilibrium (on a second-degree unrelated subset of 3,969 individuals, as determined by KING^39^) P value less than 10^−6^ and those with allele balance less than 0.3 or greater than 0.7 were also removed.

Sample-level quality control was also undertaken on this dataset; 13 samples were identified for exclusion because of singleton counts that were at least eight median absolute deviations above the median. Separately, 12 sex-discordant samples were flagged using `plink --check-sex`, and after examining chromosome Y missingness and F coefficient values for these samples, only the one that clearly differed from its reported sex was marked for exclusion. No samples were excluded based on missingness fraction or the first five principal components. In total, 14 samples were excluded from the heritability, GWAS, and Mendelian randomization analyses; the other analyses were performed without exclusion of these samples. As a result, the former analyses were performed with N = 4,149 while the latter had N = 4,163.

### Mitochondrial genome copy number estimation

We estimated mitochondrial genome copy number (MT-CN) from both WGS and WES data. In WGS data, we used BEDTools^40^ to calculate per-base coverage on the mitochondrial genome from the latest available 4,163 WGS CRAM files. MT-CN was then calculated by normalizing the mean coverage of the mitochondrial genome to the “haploid coverage” of the autosomes as calculated by Picard^32^. The result was then doubled to account for the diploidy of the autosomal genome. This normalization is summarized by the following equation: 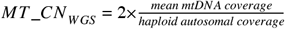

The output from the above equation served as the raw measurement of per sample MT-CN. To reduce batch effects, we separated the 4,163 samples into three groups: METSIM, FINRISK collected in 1992 or 1997, and FINRISK collected in 2002 or 2007 (the FINRISK batching decisions were made based on the means shown in **Figure S4**). Within each cohort, the raw estimates were regressed on age, age^2^ and sex (FINRISK only) and the residuals were inverse-normal transformed. We combined the three batches of normalized MT-CN values and inverse-normal transformed the combined values for downstream analysis.

We used a similar procedure to estimate MT-CN from WES data, with mean autosomal coverage estimates taken from XHMM^41^. However, as mitochondrial genomic coverage was nonuniform due to the use of hybrid capture probes, mean mtDNA coverage was not an obvious choice of metric for MT-CN estimation (**Figure S5**). To summarize this nonuniform mitochondrial genomic coverage into a single number, we tried taking the mean and the maximum depth of reads that aligned to the mitochondrial chromosome; the resulting values were then processed in the same way as the WGS-estimated values. We evaluated the approaches by measuring the R^2^ between WGS-estimated and WES-estimated MT-CN in the 4,013 samples for which both data types were available (**Figure S6**). While R^2^ was fairly high using both approaches, the maximum coverage method was ultimately selected for use as it yielded a higher R^2^ (0.445 vs 0.380). As a result, the WES MT-CN estimate was calculated as follows: 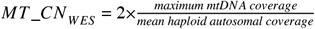.

### Mitochondrial haplogroup estimation

We assigned mitochondrial haplogroups using HaploGrep^42^ v1.0. Mitochondrial SNP/indel variants were genotyped using GATK GenotypeGVCFs, and a customized filter based on allele balance was applied to the combined callset. HaploGrep was then used to call mitochondrial haplotypes for each individual. We adjusted for major haplogroups in the same linear regressions of metabolic traits onto MT-CN (see Results) and calculated the summary statistics from a permutation test as implemented in the R package lmPerm.

### Heritability analysis

To estimate heritability of MT-CN, a genomic relatedness-based restricted maximum-likelihood (GREML) method was used as implemented in GCTA^43^. The original GREML^44^ method was used first, followed by GREML-LDMS^45^ to account for biases arising from differences in minor allele frequency (MAF) spectrum or linkage disequilibrium (LD) properties between the genotyped variants and the true causal variants^46^. For both analyses, MT-CN values were normalized and residualized for sex, age, and age^2^ as described above. Heritability estimation was performed jointly and separately for METSIM and FINRISK samples using WGS and imputed array genotypes. In all cases, a minimum MAF threshold of 1% was applied. Beyond those covariates already adjusted for in the normalization process, sensitivity analyses were performed on imputed array data to determine whether heritability estimates were sensitive to inclusion of covariates. In these experiments, either cohort or FINRISK genotyping array batch were included as fixed-effect covariates in joint analyses of imputed array data; in neither case was the final heritability estimate significantly affected (h^2^ = 0.09, SE = 0.02 in both cases). In GREML-LDMS, genotypes were split into four SNP-based LD score quartiles and two MAF bins (1% > MAF > 5% and MAF > 5%), and genetic relatedness matrices (GRMs) were estimated separately for each of the eight combinations. The GREML algorithm was then run on all eight GRMs simultaneously using the first ten principal components (PCs) of the genotype matrix (as calculated by smartPCA v13050) as fixed covariates^45^. The use of GREML-LDMS over GREML also did not affect estimated heritability values (**Table 3**), suggesting that the properties of the causal variants for this trait do not lead to significant biases when using the standard GREML approach.

**Table 3.** GREML and GREML-LDMS heritability estimates for normalized MT-CN and low-density lipoprotein (LDL).

| Trait | Genotype source (phenotype source) | N<sup>a</sup> | GREML |  | GREML-LDMS<sup>b</sup> |  |
| --- | --- | --- | --- | --- | --- | --- |
|  |  |  | h<sup>2</sup> | SE | h<sup>2</sup> | SE |
| Normalized MT-CN | WGS (WGS-measured MT-CN) | 3065 | 0.31 | 0.07 | 0.31 | 0.09 |
|  | Imputed array<sup>c</sup> (WES-measured MT-CN) | 6789 | 0.11 | 0.04 | 0.14 | 0.05 |
| LDL | WGS | 3062 | 0.34 | 0.08 | 0.38 | 0.10 |
|  | Imputed array<sup>c</sup> | 6787 | 0.25 | 0.04 | 0.32 | 0.05 |

<sup>a</sup>All analyses are limited to METSIM data
<sup>b</sup>GREML-LDMS heritability estimates are calculated using PCs 1-10 as fixed-effect covariates.
<sup>c</sup>Analyses of imputed array data exclude samples with WGS data

We observed that WGS heritability estimates decrease when analyzing FINRISK and METSIM data together compared to analysis of METSIM alone (**Table 2**) (note that FINRISK-only heritability estimates are not reliable as they have large standard errors resulting from the small number of FINRISK samples sequenced). One potential explanation for this is that there exists substantial heterogeneity across FINRISK survey years (**Figure S4**), and between the FINRISK and METSIM cohorts, with respect to the reliability with which mtDNA was captured (likely due to different DNA preparation protocols).

**Table 2.** GREML heritability estimates in each cohort separately and in joint analysis.

|  |  | WGS-measured MT-CN |  |  | WES-measured MT-CN |  |  |
| --- | --- | --- | --- | --- | --- | --- | --- |
|  |  | N<sup>a</sup> | h<sup>2</sup> | SE | N<sup>a</sup> | h<sup>2</sup> | SE |
| Joint Analysis | WGS genotypes | 4149 | 0.17 | 0.06 | 3916 | 0.11 | 0.06 |
|  | Imputed genotypes | 3916 | 0.16 | 0.06 | 17718 | 0.09 | 0.02 |
| METSIM | WGS genotypes | 3065 | 0.31 | 0.07 | 2974 | 0.20 | 0.08 |
|  | Imputed genotypes | 2974 | 0.27 | 0.08 | 9791 | 0.11 | 0.03 |
| FINRISK | WGS genotypes | 1084 | 0.20 | 0.22 | 942 | 0.24 | 0.27 |
|  | Imputed genotypes | 942 | 0.35 | 0.27 | 7927 | 0.08 | 0.03 |

<sup>a</sup>All analyses in this table were limited to sample sets with available imputed genotype data, yielding slightly lower sample sizes than in other tables.

### Genome-wide association analyses

Genome-wide association studies (GWAS) were performed using the same normalized phenotype used in heritability analyses. Single-variant GWAS were conducted using EMMAX as implemented in EPACTS. Kinship matrices required by EMMAX were generated by EPACTS; kinship matrices for WGS GWAS were generated from WGS data, while those for WES and imputed array based GWAS were generated from WES data. A P value threshold of 5×10^−8^ was used for the WGS and imputed array GWAS while 5×10^−7^ was used for significance in the WES GWAS. Single-variant association analyses of WGS and WES data did not include any covariates in the EMMAX model, although all association analyses were performed using MT-CN values that adjusted for age, age^2^, sex, and cohort (see “Mitochondrial Genome Copy Number Estimation”). All association tests labeled “joint” were performed on METSIM and FINRISK cohorts together; in one case, a random-effects meta-analysis was performed using individual-cohort summary statistics and the R package meta^47^.

Gene-based variant aggregation studies (RVAS) were done using a mixed-model version of SKAT-O^48^ as implemented in EPACTS. Variants with CADD^49^ score greater than 20 and minor allele frequency less than 1% were grouped into genes as annotated by VEP^50^ (which annotates a variant with a gene name if the gene falls within 5k of that gene by default). For gene-based RVAS, genome-wide significance thresholds varied slightly due to differing the number of genes with at least two variants meeting the above criteria in each test, but were approximately 2×10^−6^ in all cases.

### Mendelian randomization

To assess the evidence for a causal relationship between mitochondrial genome copy number and fasting serum insulin levels, the METSIM cohort alone was used due to its homogeneity of sex, collection procedures, and location. A penalized regression based, multiple variant Mendelian randomization (MR) approach was employed to enforce the necessary assumptions of MR methods. While some MR studies have tested one or more assumptions *post hoc*, to our knowledge, there is no published method that tries to enforce these assumptions during the process of building the genetic instrument in the absence of a large set of known genotype-exposure associations. In our formulation (**Figure 4a**), X, the natural log of MT-CN (adjusted for nuclear genomic coverage but not for age, age^2^, or sex), and a genotype matrix G were used to build a genetic instrument Z, which was then tested against Y, the natural log of fasting serum insulin. The goal of the MR approach was to use a large number of common variants to build a genetic instrument Z that satisfied the three assumptions of MR^51^:

**Figure 4.**
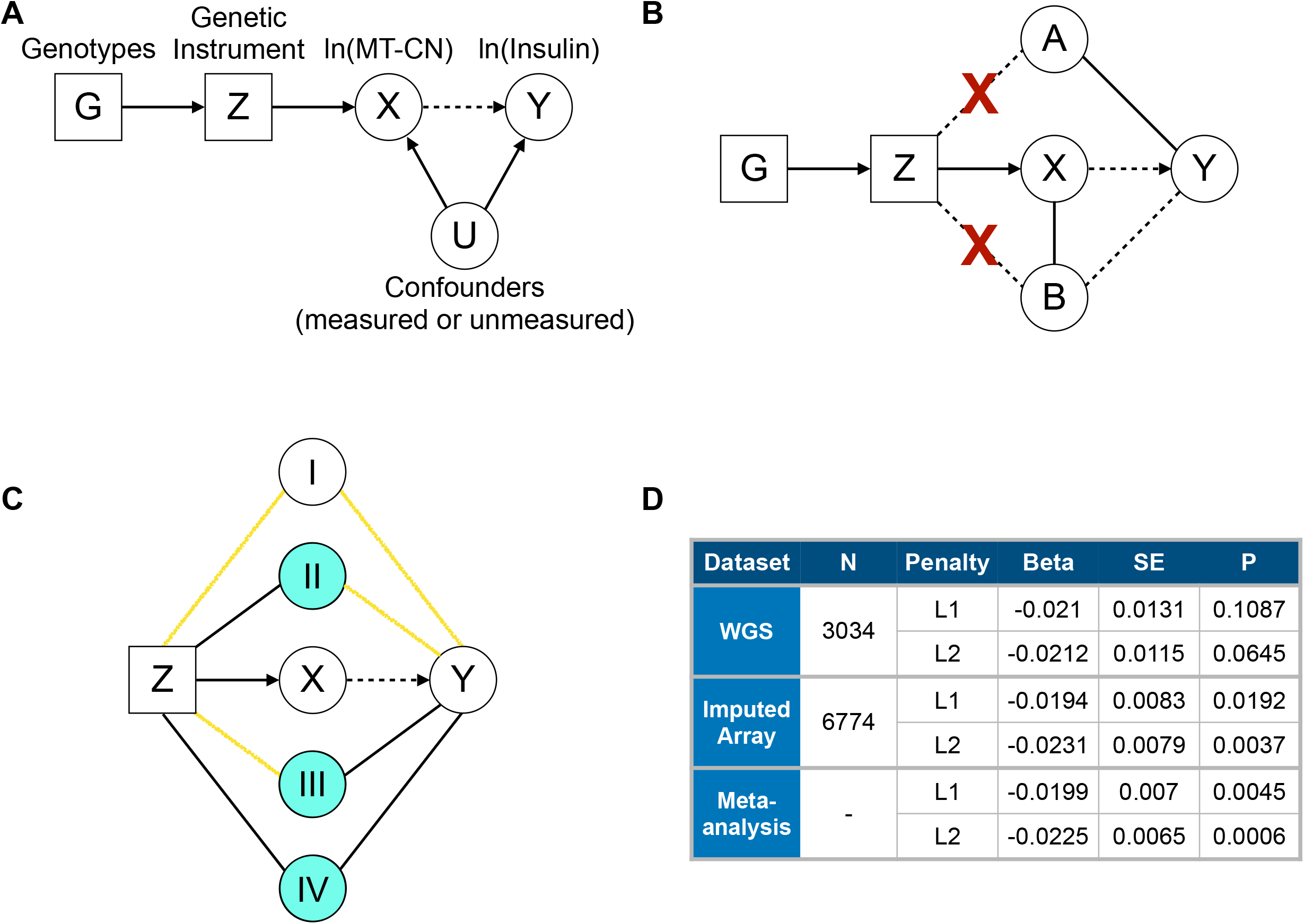
Mendelian randomization approach and results. (A) Formulation of the Mendelian Randomization causality test. G represents genotypes, Z is a genetic instrument value constructed from G, X represents ln(MT-CN), Y represents ln(Insulin), and U represents any confounders of the association between X and Y. The arrow from X to Y is dashed to indicate that although an association is known, the relationship is not known to be causal. In this formulation, a significant association between Z and Y would provide evidence that X is casual for Y. (B) Strategy for choosing variables to adjust for when building Z in order to enforce MR assumptions. A represents those columns of covariate matrix W that are associated with Y (represented by the solid line between A and Y) and B represents those columns of matrix W that are associated with X conditional on A (represented by the solid line between B and X). Dashed lines represent possible, but unproven associations. The penalized regression of X on G used to build Z is adjusted for A and B (with no penalty) in an attempt to prevent any associations between Z and either A or B (represented by the blue X’s). While an association between B and Y is unlikely (represented by the dashed line between B and Y) because B is not contained in A, B is still adjusted for in the penalized regression to be as conservative as possible. (C) Strategy for choosing covariates to adjust for in the causality test of Y against Z in another attempt to reduce the impact of any remaining associations between Z and assumption-violating variables. Covariate sets I, II, III, and IV are defined by the presence of known first-order associations (represented by black lines) with Z and Y (see Material and Methods). Yellow lines represent relationships where a first-order association is not known, but a higher-order association is possible. Covariate sets II, III, and IV (colored blue) are adjusted for in the causality test because there is at least one first order association linking them to Z and Y, so they risk violating MR assumptions 2 or 3. (D) Results of Mendelian randomization test for causality of MT-CN on fasting serum insulin.

1. Association of Z with X
2. Independence of Z from any variables U confounding the relationship between X and Y
3. Independence of Z and Y given X and U

To attempt to build a genetic instrument satisfying assumptions 2 and 3, the deep METSIM phenotype data were leveraged. A matrix W was constructed using the 75 measured traits and first 20 PCs of the genotype matrix (including a third-degree polynomial basis for PC 1). From these variables, covariates that could violate one of these two assumptions were chosen by selecting columns of W associated with X or Y (**Figure 4b**). These columns were selected using two successive LASSO feature selection procedures. First, a set A of covariates associated with Y was chosen by using LASSO to regress Y onto W. In this regression, age and the third-degree polynomial basis for PC 1 were left unpenalized to ensure that A contains these covariates. The shrinkage parameter was chosen by tenfold cross-validation as the largest value that gives a mean squared error (MSE) within one standard error of the minimum observed MSE. Next, the columns of W associated with X conditional on A were chosen using a similar LASSO procedure in the regression of X onto W. In this step, however, the variables in set A were left unpenalized in order to only capture associations that are conditionally independent of A. The selected variables from this regression were designated set B.

The instrument was built using a penalized regression (using either an L1 or L2 penalty, as implemented in glmnet^52^) of the form *X* ∼ *G* + *W _A_* + *W*_*B*_, where W_A_ and W_B_ are the columns of W representing sets A and B, respectively, and G is a genotype matrix containing the alternate allele dosage (missing alleles are replaced with the MAF, similarly to PLINK^53^) of all variants with MAF greater than 1% and marginal GWAS P value below 0.01. As X was the target vector for this regression, assumption 1 of MR was trivial. In the penalized regression, W_A_ and W_B_ were unpenalized in an effort to orthogonalize the regression coefficients of the genotypes to these covariates in an effort to enforce assumptions 2 and 3. glmnet was run with a convergence threshold of 1×10^−10^ and maximum number of iterations of 200,000. To avoid the overfitting that would result from calculating instrument values on the same samples on which regression coefficients are learned^54^, the penalized regression model was fit on independent subsets of the data as follows. Five models were fit, each by holding out a different 20% of samples, such that the instrument value computed for each sample was calculated using the regression coefficient vector learned without that sample. The vector of possible shrinkage parameters λ for all five models was supplied as (10^3^,10^2^,…,10^−13^,10^−14^), and the λ value which minimized the joint residual sum of squares of all five models was chosen for instrument calculation.

Formally, we randomly partitioned the set of samples S with nonmissing insulin measurements into five nonoverlapping sets S_j_ for *j* = {1, …, 5}. We denote set complements as 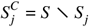, such that each S_j_^c^ contained 80% of the training samples. The instrument vector Z_j_ for each S_j_ was computed as follows: 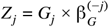, where Z_j_ is the instrument vector for S_j_, G_j_ is the genotype matrix of S_j_, and β_G_ ^(-j)^ is the vector of genotype regression coefficients from the model described above, trained on S_j_ ^C^. The instrument values within each S_j_ were inverse rank-normalized using a Blom transformation^55,56^ before being concatenated across the values of j to give the final instrument vector Z. Because samples with missing insulin values could not be included in the causality test anyway, these samples were excluded from S but safely included in the training sets of all five models. The instrument values of these samples were never calculated or used in downstream analyses.

Often, the inclusion of unpenalized covariate sets A and B in the instrument-building regression was not sufficient to completely orthogonalize Z to these covariates (see below). As a result, the test for association between Z and Y was performed conditional on a set of potentially assumption-violating covariates chosen using the newly constructed instrument Z in another attempt to account for possible violations of MR assumptions in the causality test (**Figure 4c**). To choose this set of covariates C, a final feature selection step was performed using LASSO regression of Z on W with covariate set A excluded from the penalty. As in the previous feature selection steps, the shrinkage parameter was chosen via tenfold cross-validation as the largest value with MSE within one standard error of the minimum observed MSE. Once this set, D, of covariates associated with Z was chosen, the covariates in W were partitioned into sets I, II, III, and IV based on their membership in A and D (see **Figure 4**). Formally, this partitioning was done as follows: *I* = *W* | (*A* ⋃ *D*), *II* = *D* ⋂ *A*^*C*^, *III* = *A* ⋂ *D*^*C*^, and *IV* = *A* ⋂ *D*, where *A*^*C*^ = *W* \ *A* and *D*^*C*^ = *W* \ *D*. Then, the test for causality came from the regression coefficient of Z in the multiple regression *Y* ∼ *Z* + *C*, where C is the union of sets II, III, and IV (colored blue in **Figure 4c**).

To account for missing data in W, missing values were multiply imputed using regression trees as implemented in the R package mice^57^ v3.4.0 `maxit=25`). This imputation was repeated 1000 times in parallel, with each set of imputed values being carried through the entire procedure described above. The resulting 1000 computed instrument effect sizes and standard errors were combined using Rubin’s method as implemented in the R package Amelia^58^ v1.7.5. The combined effect size and standard error were then tested for significance using a t-test with 998 degrees of freedom.

The above procedure was performed separately for METSIM samples with WGS data (N = 3,034) and METSIM samples with only imputed array data (N = 6,774) using an L1 penalty in the instrument-building regression, and again using an L2 penalty. Both sample sets were limited to those for which relevant quantitative traits were available. An inverse-variance weighted meta-analysis was performed across data sets for L1 and L2-penalized regression separately. The resulting effect size and standard error were tested for significance using a Z test.

To ensure that our results were not driven by outlier samples, we removed outliers in two stages. Before the MR analyses, we used principal components analysis (PCA), Mahalanobis distance, and multi-trait extreme outlier identification to remove 5 WGS samples and 15 imputed array samples based on quantitative trait data. We also removed high leverage, high residual outliers from the causality test regression (see below) *post hoc* and recomputed the instrument effect sizes to ensure that there was no significant change in the results. In each of the 1,000 multiple imputation runs, among the samples with standardized residual greater than 1, the top 10 samples by leverage were recorded. Any sample that was recorded in this way in at least one run was then excluded from the re-analysis as a *post hoc* outlier. The results of this additional analysis showed only very small differences in effect estimates, and their interpretation remained the same (**Table S9**). Thus, we concluded that our causal inference results were not driven by outlier samples.

One caveat of this method is that, as mentioned above, exclusion of sets A and B from the regression penalty did not perfectly orthogonalize the resulting instrument from these variables in practice (**Figure S7**). Reasons for this may include relatively low levels of shrinkage in the instrument-building regression or higher order associations between MT-CN and the confounding variables. However, our method still represents an improvement over the current standard, which is not to adjust for these covariates at all. Another caveat is that it is impossible to determine the perfect set of covariates for which adjustment is appropriate. Lack of adjustment for truly confounding variables can result in an instrument which does not satisfy MR assumptions 2 and/or 3, yielding a biased effect estimate. Conversely, unnecessary adjustment for certain variables can also result in biases. For example, adjusting for an intermediate phenotype that truly lies along the path from Z to X to Y can cause a false negative signal, making the causality test overly conservative. Alternatively, adjusting for some variables can result in collider biases^59^. That is, if both Z and Y are causal for a confounder U, then adjusting for U can induce a dependency between Z and Y (**Figure S8**) that did not previously exist.

We note that a known source of bias in MR studies is the selection of samples based on case-control status for a related disease^60^. While METSIM is a population-based study, samples were selected for WGS based on cardiovascular disease case-control status so as to enrich the sequenced samples for cases. This has the potential to bias a MR experiment if both the exposure and the outcome are associated with the disease, which is certainly possible. However, in our design, all of the METSIM samples not chosen for WGS were tested in the imputed array experiment. The consistency of effect estimates between the WGS and imputed array samples both in the L1 and L2 penalty cases (**Figure 4d**) suggests that there is little to no bias arising from sample selection in this experiment.

### Calculation and testing of polygenic risk score in the UK Biobank

To search for associations between MT-CN and other phenotypes, the genetic instrument calculated in Finnish imputed array data was computed and treated as a polygenic risk score (PRS) in a relatively homogenous subset of 357,656 UK Biobank samples identified by a previous study^61^. We calculated 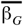, the average of the five values of β_G_ ^(-j)^ across all 1000 multiple imputation runs using an L2 penalty and imputed array data – the L2 penalty was chosen because it performed better than the L1 on both METSIM data types, and the imputed array data set was chosen due to its larger sample size than the WGS set (**Figure 4d**). Next, to keep the procedure as consistent as possible with the imputation protocol used for METSIM – which used haploid dosage values to call imputed genotypes^36^ – we called imputed genotypes using the expected alternate allele dosage from the UK Biobank by setting thresholds of 0.5 and 1.5. Using the resulting imputed variant calls, we calculated our PRS as 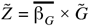, where 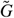 is the UK Biobank genotype dosage matrix constructed in the same way as G in METSIM.

To test for associations with MT-CN PRS in the UK Biobank, we employed two approaches: a hypothesis-driven analysis targeted to the phenotypes associated with MT-CN in the Finnish data as well as a hypothesis-free screen of all the phenotypes available to us.

In the targeted analysis, we used our genetic instrument from the MR experiment as a PRS for MT-CN in our chosen subset of the UK Biobank and tested for associations with several blood cell count and metabolic syndrome traits. Given the association of MT-CN with rs9389268 (see Results), we selected as cell count traits total leukocyte count as well as lymphocyte, neutrophil, monocyte, and platelet counts for testing (because lymphocyte count was not readily available, it was calculated as the product of leukocyte count and lymphocyte percentage). We did not include basophils and eosinophils in this analysis considering that they comprise a small minority of white blood cells and are unlikely to affect MT-CN measured from whole blood. All cell count traits were log-transformed and standardized separately by sex.

We took several steps to eliminate outlier samples in the dataset. Through three iterations of PCA on the cell count matrix and subsequent outlier removal, we removed 1,637 outlier samples. We then fit null linear models of the form *cell count* ∼ *age* + *age*^2^ + *sex* + *age* : *sex* + *age*^2^ : *sex* + *PCs* (the first 20 PCs were included) for each cell count trait and subsequently removed samples with either large residuals or high leverage and moderate residuals in at least one model (following the example of ^61^). Through two iterations of null model fitting and outlier removal, we removed 7 additional samples based on null model fit. Tests of association between cell counts and MT-CN PRS were based on the PRS regression coefficient in linear models of the form *cell count* ∼ *PRS* + *age* + *age*^2^ + *sex* + *age* : *sex* + *age*^2^ : *sex* + *PCs*.

We repeated this process for those cardiometabolic traits found to be suggestively associated with MT-CN in the Finnish dataset (P < 10^−6^) that were also readily available in the UK Biobank (**Figure 1a, Table S1**); these phenotypes were body mass index (BMI), fat mass, C-reactive protein, high-density lipoprotein, total triglycerides, and weight. We also chose to include T2D status because of the lack of insulin measurement in the UK Biobank. Except for T2D, a binary trait, all traits were log-transformed before further analysis (after removing 817 samples with negative values for T2D, representing missing information). The above outlier removal steps were repeated for the cardiometabolic traits after excluding the outliers already identified from the cell count data, with the only major modification being the use of logistic regression for the T2D models. This process resulted in the removal of 42 and 53 samples from PCA and null model fitting, respectively.

**Figure 1.**
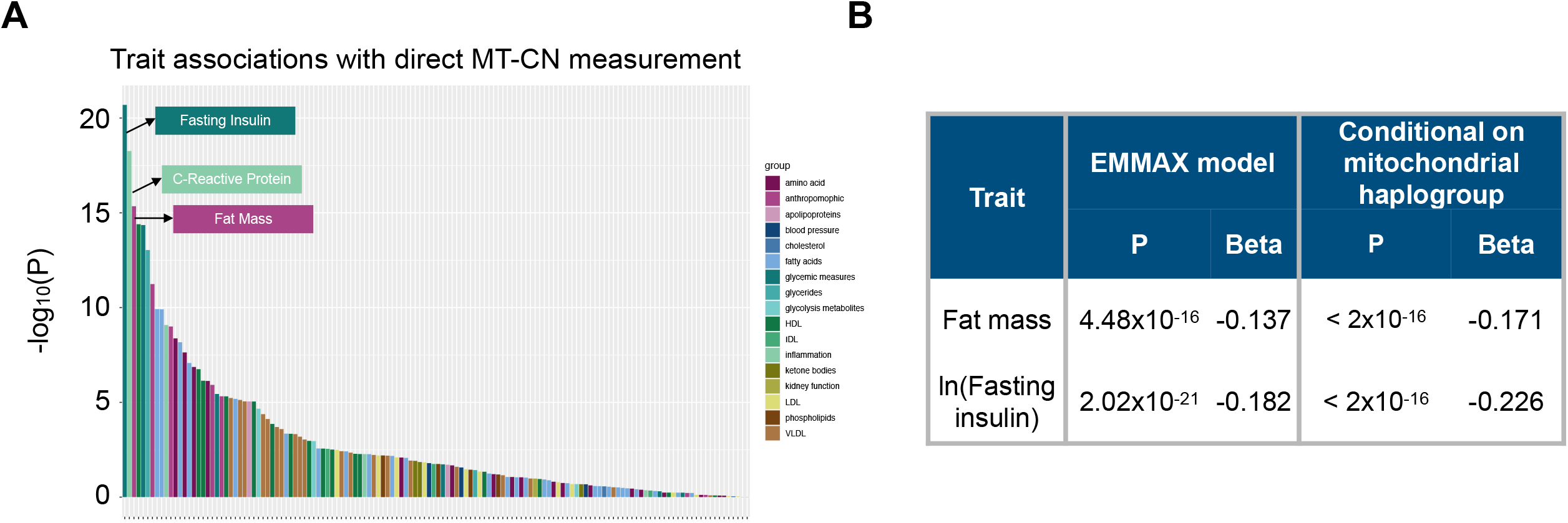
Cardiometabolic trait associations with MT-CN in WGS data. (A) Phenome-wide association study of normalized MT-CN against 137 cardiometabolic traits in the 4,163 sample data set. Traits are grouped into 17 categories, represented by the color of each bar. The top three most significant traits are, in order: fasting serum insulin, C-reactive protein, and fat mass. Exact P values and effect estimates, as calculated by EMMAX, are listed in **Table S1**. (B) Association tests between normalized MT-CN and both fat mass and fasting serum insulin using WGS data (N = 4,163). Results are shown for the EMMAX test and a permutation test in which mitochondrial haplogroups were adjusted for.

SKAT-O tests of association between *TMBIM1* and the cell count traits identified above were also performed. Similarly to the RVAS in Finnish data, variants within 5kb of *TMBIM1* with MAF < 1% and CADD v1.6 score > 20 were selected for inclusion in this analysis. Rather than the mixed-model version of SKAT-O used in the Finnish data, standard SKAT-O was used due to the lower expected level of cryptic relatedness in the UK Biobank population.

We also performed a hypothesis-free, phenome-wide screen of UK Biobank traits to which we had access (**Table S10**), to search for other associations with MT-CN PRS. The statistical models used in this screen were of the same form as those described above, both with and without adjustment for neutrophil and platelet counts. To curate and transform phenotypes, we used an adapted version of PHESANT^61,62^. A few further modifications were made to the pipeline, the most significant being the direct use of logistic regression for testing categorical unordered variables, the inclusion of cancer phenotypes, and the exclusion of sex-specific (or nearly sex-specific) categorical traits. The PHESANT pipeline we used^61^ outputs continuous variables both in their raw form and after applying an inverse rank normal transformation. For the sake of being conservative and robust to outliers, we chose to interpret the results from the normalized continuous variables. To control false discovery rate, we performed a Benjamini-Hochberg procedure with Storey correction as implemented in the R package qvalue^63^ v2.18.0 on the categorical and normalized continuous variables together. As a secondary analysis, this same correction was applied to the categorical and raw continuous variables together.

## Results

### Association of MT-CN with metabolic traits

We estimated MT-CN in 4,163 individuals from the METSIM and FINRISK studies based on deep (>20x coverage) WGS data. We did so by measuring the mean coverage depth of reads mapped to the mitochondrial genome in each sample, and normalizing it to the mean autosomal coverage (see Material and Methods). We performed batch normalization separately for METSIM and for two FINRISK batches separated by survey years (see Material and Methods). Each measurement was adjusted for age, age^2^, and sex, then inverse rank normalized separately before combining across batches. We tested the resulting MT-CN estimates for association with 137 quantitative traits that were collected and normalized according to the procedures described previously^36^. MT-CN was strongly associated with fat mass (P = 4.48 × 10^−16^) and fasting serum insulin (P = 2.02 × 10^−21^), as well as numerous additional quantitative traits, many related to metabolic syndrome **(Figure 1a, Table S1**). Notably, BMI was significantly associated with MT-CN, although Ding *et al*. did not find evidence of this association^11^. Since population structure was a potential confounder in this analysis considering the presence of mtDNA polymorphisms that might adversely affect short-read alignment, we included SNP-inferred mitochondrial haplogroup as a covariate and reran the tests (**Figure 1b**). The association signals retained significance even after this adjustment.

To understand the connection between MT-CN and more clinically relevant phenotypes, we tested our MT-CN estimate against Matsuda ISI and disposition index (**Table 1**), which measure insulin sensitivity and secretion, respectively, and were not included in the initial screen. MT-CN was strongly associated with both insulin phenotypes. Notably, the Matsuda ISI signals survived adjustment for fat mass percentage after excluding diabetic individuals, which indicates that the association of peripheral blood MT-CN with insulin sensitivity was independent of fat mass.

**Table 1.**
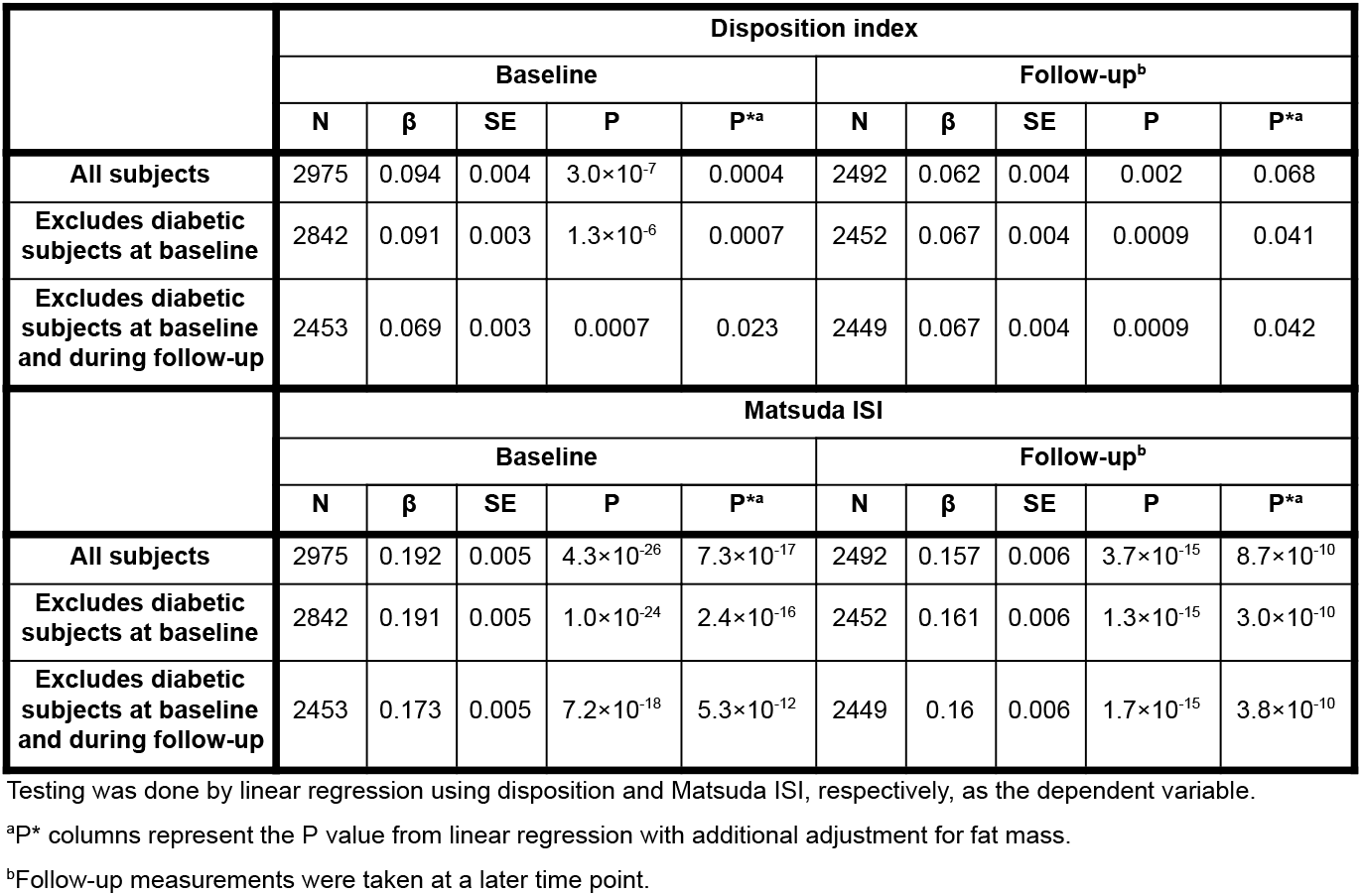
Associations of normalized MT-CN with disposition index and Matsuda ISI in METSIM.

|  | Disposition index |  |  |  |  |  |  |  |  |  |
| --- | --- | --- | --- | --- | --- | --- | --- | --- | --- | --- |
|  | Baseline |  |  |  |  | Follow-up<sup>b</sup> |  |  |  |  |
|  | N | β | SE | P | P<sup>*a</sup> | N | β | SE | P | P<sup>*a</sup> |
| All subjects | 2975 | 0.094 | 0.004 | 3.0×10<sup>-7</sup> | 0.0004 | 2492 | 0.062 | 0.004 | 0.002 | 0.068 |
| Excludes diabetic subjects at baseline | 2842 | 0.091 | 0.003 | 1.3×10<sup>-6</sup> | 0.0007 | 2452 | 0.067 | 0.004 | 0.0009 | 0.041 |
| Excludes diabetic subjects at baseline and during follow-up | 2453 | 0.069 | 0.003 | 0.0007 | 0.023 | 2449 | 0.067 | 0.004 | 0.0009 | 0.042 |
|  | Matsuda ISI |  |  |  |  |  |  |  |  |  |
|  | Baseline |  |  |  |  | Follow-up<sup>b</sup> |  |  |  |  |
|  | N | β | SE | P | P<sup>*a</sup> | N | β | SE | P | P<sup>*a</sup> |
| All subjects | 2975 | 0.192 | 0.005 | 4.3×10<sup>-26</sup> | 7.3×10<sup>-17</sup> | 2492 | 0.157 | 0.006 | 3.7×10<sup>-15</sup> | 8.7×10<sup>-10</sup> |
| Excludes diabetic subjects at baseline | 2842 | 0.191 | 0.005 | 1.0×10<sup>-24</sup> | 2.4×10<sup>-16</sup> | 2452 | 0.161 | 0.006 | 1.3×10<sup>-15</sup> | 3.0×10<sup>-10</sup> |
| Excludes diabetic subjects at baseline and during follow-up | 2453 | 0.173 | 0.005 | 7.2×10<sup>-18</sup> | 5.3×10<sup>-12</sup> | 2449 | 0.16 | 0.006 | 1.7×10<sup>-15</sup> | 3.8×10<sup>-10</sup> |

Testing was done by linear regression using disposition and Matsuda ISI, respectively, as the dependent variable.
<sup>a</sup>P\* columns represent the P value from linear regression with additional adjustment for fat mass.
<sup>b</sup>Follow-up measurements were taken at a later time point.

To test for this association signal in a larger cohort, we developed a method to estimate mitochondrial genome copy number using 19,034 samples with whole exome sequencing (WES) data from the METSIM and FINRISK studies that included most of the WGS samples^36^ (see Material and Methods). R^2^ between WGS-based and WES-based estimates was 0.445 (**Figure S6**). Consistent with the WGS-based analysis, WES-estimated MT-CN was significantly associated with both fat mass and fasting serum insulin levels, even after removing the samples with WGS data, with identical directions of effect (**Table S2**).

Anecdotally, it is interesting to note that these MT association signals can also be detected using read-depth analysis of the nuclear genome (**Figure S1;** manuscript under review^64^ - preprint doi: 10.1101/2020.12.13.422502), where reads derived from mtDNA align erroneously to several nuclear loci based on homology between the MT genome and ancient nuclear mitochondrial insertions. This result provides additional evidence for the reported trait associations using an independent MT-CN estimation method, and indicates that these homology-based signals need to be taken into account in future CNV association studies.

### Heritability analysis

To assess the extent to which MT-CN is genetically determined, we estimated the heritability of mitochondrial genome copy number using GREML (**Table 2**). We explored two different approaches available: (1) analysis of the 4,149 samples with WGS data that passed quality control measures, where both nuclear genotypes and MT-CN are measured directly from the WGS data, and (2) analysis of the set of 17,718 samples with imputed genotype array data, where MT-CN is estimated from WES data. Of these, (1) benefited from more accurate measurement of genotype and phenotype, whereas (2) had noisier measurements but benefited from larger sample size. We focused primarily on the METSIM cohort, both because of the homogeneity of this cohort (see Material and Methods) and because the number of FINRISK samples with WGS data was small.

In the WGS analysis, the GREML-estimated heritability of MT-CN in METSIM was 31% - somewhat less than the 54% value reported in the only prior large-scale study of peripheral blood MT-CN heritability, which was based on low-coverage WGS^11^. For comparison, we used this same approach to estimate heritability of LDL in METSIM WGS data, which yielded an estimate of 34% with a standard error of 7.9% (**Table 3**). This is broadly consistent with prior work^65,66^, including analysis of the same Finnish sample set using distinct methods^36^ (20.2% heritability). These results show that mitochondrial genome copy number is a genetically determined trait with significant heritability, comparable to that of LDL and other quantitative cardiometabolic traits^36^.

The analysis of imputed METSIM genotypes using WES-estimated MT-CN yielded an estimated heritability of 11%, which is much lower than the WGS-based estimate (**Table 2**). To understand this discrepancy, we repeated the GREML analysis with the other two combinations of phenotype source (WGS vs. WES estimation) and genotype source (WGS vs. imputed array). When using the WGS-measured phenotype, the estimated heritability decreased only slightly (31% to 27%) when switching from the WGS to imputed genotypes. This suggests that the difference in genotyping method was not the main driver of the observed heritability disparity between the WGS and imputed array datasets. Conversely, when analyzing the imputed METSIM genotypes, switching from WGS-measured to WES-measured MT-CN resulted in a large drop (27% to 11%) in estimated heritability. This suggests that the extra noise inherent in WES-based MT-CN estimates was responsible for the reduction in the GREML-estimated heritability despite the increased sample size of the imputed array dataset.

### Identification of genetic factors associated with MT-CN

Previous studies have identified three autosomal quantitative trait loci (QTL) reaching genome-wide significance for MT-CN in other populations^25,26^. Another recent study identified two putative QTLs with suggestive P values^27^. We conducted single variant GWAS for MT-CN (see Material and Methods). Analysis of WGS (N = 4,149) and WES (N = 19,034) genotypes yielded no variants exceeding the respective significance thresholds of 5×10^−8^ and 5×10^−7^ (**Figure S2**). However, despite the increased noise in the WES-measured phenotype, GWAS of imputed array genotypes from METSIM (N = 9,791) yielded two loci with genome-wide significant associations, identified by lead markers rs2288464 and rs9389268 (**Figure 2, Table 4**). Of the previously-reported MT-CN QTLs^25–27^, we observed an inconclusive signal at rs445 (P = 0.048) and a significant signal at rs709591 (P = 1.61×10^−4^), a locus associated with neutrophil count^67,68^ (**Table S11**). No significant signal was observed at the other two single-variant QTLs (**Table S11**) or the linkage peak identified by Curran *et al*. (**Figure S3**).

**Table 4.** Single marker association results for rs2288464 and rs9389268.

|  |  | FINRISK<sup>a</sup> |  |  |  | METSIM |  |  |  | Joint Analysis |  |  |  |
| --- | --- | --- | --- | --- | --- | --- | --- | --- | --- | --- | --- | --- | --- |
|  |  | N | MAF | P<sup>b</sup> | Beta | N | MAF | P<sup>b</sup> | Beta | N | MAF | P<sup>b</sup> | Beta |
| rs2288464 | Imputed | 7927 | 0.148 | 0.613 | 0.0113 | 9791 | 0.165 | *2.55×10<sup>-9</sup>* | 0.119 | 17718 | 0.158 | *9.77×10<sup>-7</sup>* | 0.075 |
|  | WES | 9221 | 0.150 | 0.376 | 0.0186 | 9813 | 0.166 | *6.75×10<sup>-9</sup>* | 0.118 | 19034 | 0.158 | *9.34×10<sup>-7</sup>* | 0.0734 |
|  | WGS | 1084 | 0.142 | 0.383 | 0.0532 | 3065 | 0.161 | 0.113 | 0.0561 | 4149 | 0.156 | 0.0655 | 0.0562 |
| rs9389268 | Imputed | 7927 | 0.354 | 0.189 | 0.0216 | 9791 | 0.347 | *1.26×10<sup>-10</sup>* | 0.0973 | 17718 | 0.35 | *1.62×10<sup>-8</sup>* | 0.0634 |
|  | WGS | 1084 | 0.351 | 0.788 | 0.0121 | 3065 | 0.347 | *3.24×10<sup>-8</sup>* | 0.150 | 4149 | 0.348 | *7.87×10<sup>-7</sup>* | 0.115 |

<sup>a</sup>Analyses of imputed FINRISK array data were performed with covariates for FINRISK genotyping batch.
<sup>b</sup>Italicized results are significant at the appropriate threshold for the given test (see Material and Methods).

**Figure 2.**
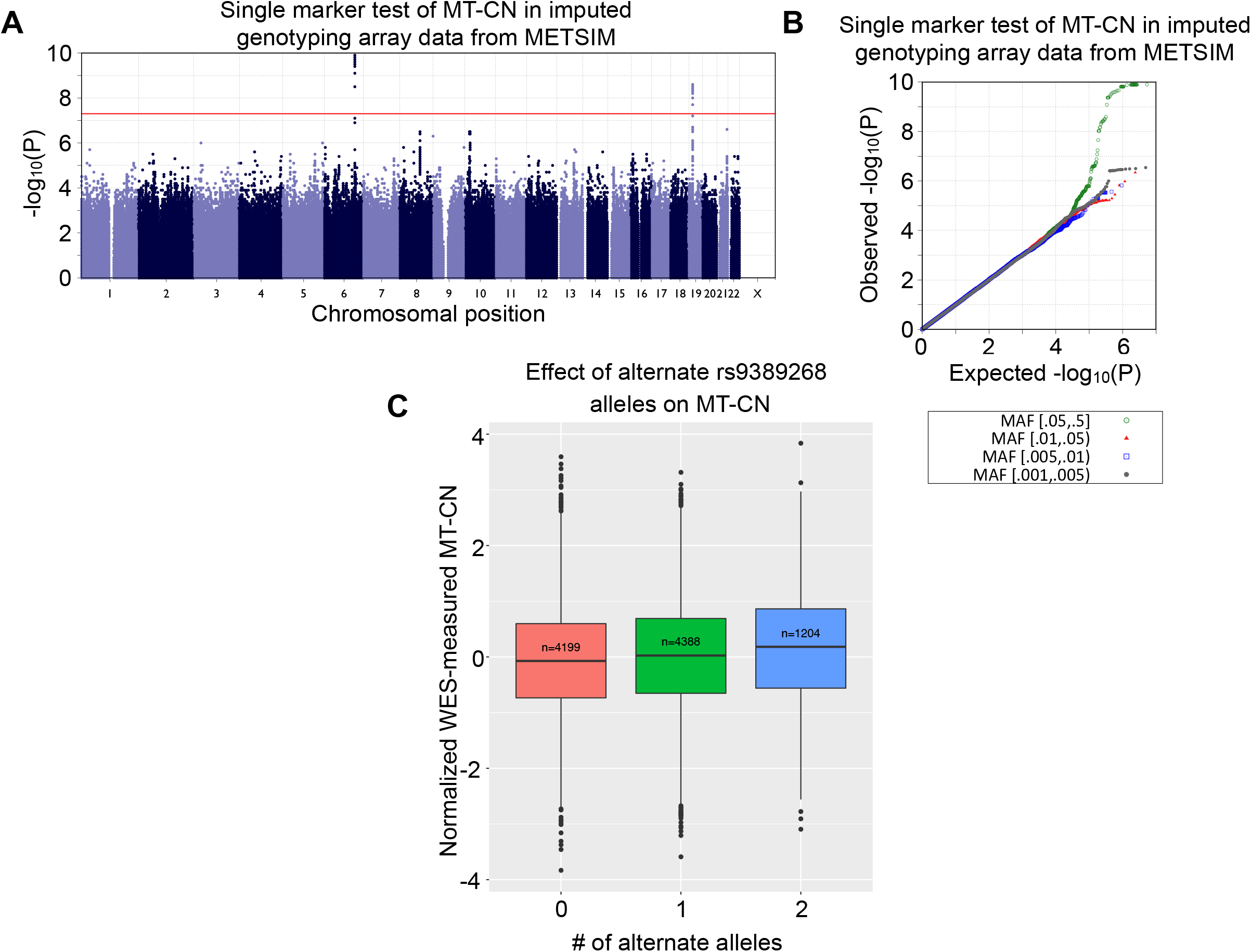
Single-marker genetic associations with MT-CN in imputed array data. (A) Manhattan plot for a genome-wide association test of normalized, WES-measured MT-CN using imputed array genotype data from METSIM (N = 9,791). Two loci markers reached the genome-wide significance of 5×10^−8^, identified by lead markers rs2288464 and rs9389268. (B) Quantile-quantile (QQ) plot for the association test shown in (A). This plot is separated by minor allele frequency bin, as indicated by the colors and shapes of the points. (C) Boxplot showing the distributions of normalized WES-measured MT-CN in METSIM separated by the number of rs9389268 alternate alleles as detected by imputed array genotyping (N = 9,791). The EMMAX P value for this variant was 1.62×10^−8^ in imputed data.

rs9389268 was the only marker that was strongly associated with MT-CN in the METSIM analyses of both WGS and imputed array data (P = 3.24×10^−8^ and P = 1.26×10^−10^, respectively). Although this variant was not significantly associated with MT-CN in FINRISK (P = 0.788 and P = 0.189 in WGS and imputed array data, respectively) or in a separate random-effects meta-analysis of both cohorts (P = 0.115), the lack of signal in FINRISK is likely the product of lower-quality MT-CN measurements in FINRISK, which displayed heterogeneity across survey years (**Figure S4**). This variant is located in an intergenic region between the *MYB* and *HBS1L* genes, is common across many populations, and is slightly more frequent in Finns compared to non-Finnish Europeans (gnomAD v3 MAF 34.4% vs. 26.0%). *MYB* and *HBS1L* are hematopoietic regulators^69,70^, and the region between them is known to be associated with many hematological parameters including fetal hemoglobin levels, hematocrit, and erythrocyte, platelet, and monocyte counts^71–74^. It has been suggested that these intergenic variants function by disrupting *MYB* transcription factor binding and disrupting enhancer-promoter looping^75^. Conditioning the METSIM-only imputed array GWAS on rs9399137 – a tag SNP shown to be associated with many of these hematological parameters^73^ – resulted in elimination of the rs9389268 signal entirely (P = 0.408), suggesting that the haplotype responsible for the association of rs9389268 with MT-CN in our data is the same one previously known to be associated with numerous hematological phenotypes.

This result is not surprising considering our approach for normalizing MT-CN. Because our MT-CN estimate was based on the ratio of mtDNA coverage to nuclear DNA coverage, changes in the cell type composition of blood could result in changes in our normalized measurement if the underlying cell types have different average numbers of mitochondria. This is especially true of platelets, which can contain mitochondria but not nuclei, and whose counts are known to be associated with rs9399137.

rs2288464 seemed to be a good candidate due to its location in the 3’ untranslated region of *MRPL34*, which codes for a large subunit protein of the mitochondrial ribosome. While the association signal at this marker was not observed in the WGS data (P = 0.0655), based on the observed effect size of this variant in WES and imputed data as well as the number of WGS datasets available, there was insufficient power (∼0.5% at α = 5×10^−7^) to robustly detect this association in the WGS data^76^.

We next performed rare variant association (RVAS) analyses using a mixed-model version of SKAT-O^48^ to test for genes in which the presence of high-impact rare variants might be associated with MT-CN levels (see Material and Methods; **Figure 3, Table 5**). Using WES data, the only gene passing the Bonferroni-adjusted P value threshold of 2.16×10^−6^ was *TMBIM1* (P = 2.96×10^−8^), a member of a gene family thought to regulate cell death pathways^77^. *TMBIM1* has been shown to be protective against non-alcoholic fatty liver disease (NAFLD), progression to non-alcoholic steatohepatitis, and insulin resistance in mice and macaques^78^. Interestingly, in our analysis - in which a burden test was determined to be optimal by SKAT-O-rare, putatively high-impact variants in *TMBIM1* were associated with a higher MT-CN (**Figure 3c**). Higher MT-CN was, in turn, associated with less severe metabolic syndrome, suggesting that *TMBIM1* is actually a risk gene, not a protective one. Thus, the published function of *TMBIM1* makes it a strong candidate, although the direction of effect in our data disagreed with the direction suggested by prior work in model organisms^78^.

**Table 5.** Gene-based rare variant association results for *TMBIM1*.

|  | FINRISK |  |  | METSIM |  |  | Joint Analysis |  |  |
| --- | --- | --- | --- | --- | --- | --- | --- | --- | --- |
|  | N | Fraction with rare allele | P | N | Fraction with rare allele | P<sup>a</sup> | N | Fraction with rare allele | P<sup>a</sup> |
| WES | 9221 | 0.016 | *1.57×10<sup>-3</sup>* | 9813 | 0.013 | *1.44×10<sup>-6</sup>* | 19034 | 0.014 | *2.96×10<sup>-8</sup>* |
| WGS | 1084 | 0.028 | 0.489 | 3065 | 0.014 | 0.01 | 4149 | 0.013 | 0.01 |

*TMBIM1* was the only genome-wide significant gene in the WES rare-variant association tests of METSIM and the whole dataset.
<sup>a</sup>Italicized results are significant at the appropriate threshold for the given test (see Material and Methods).

**Figure 3.**
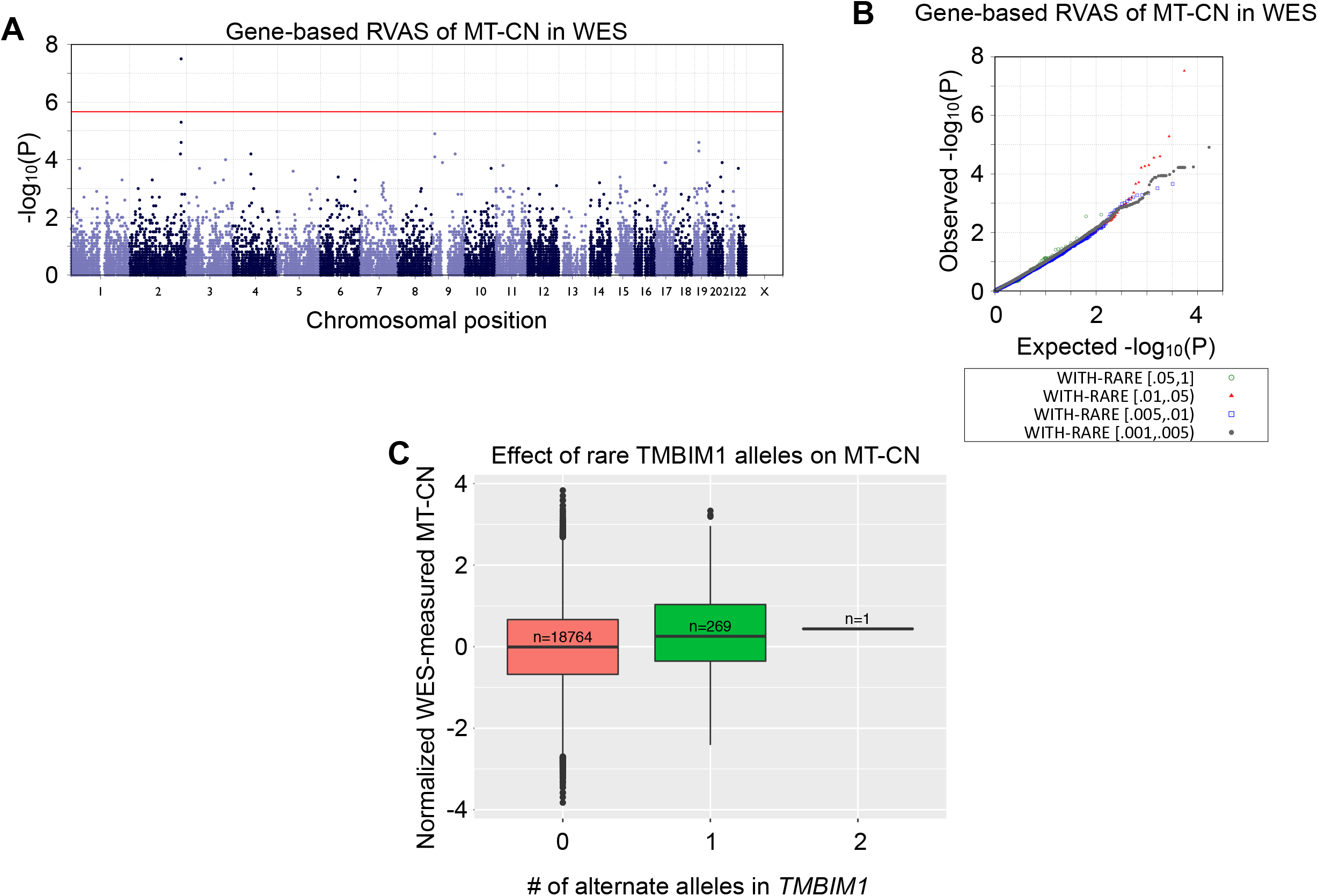
Gene-based associations with MT-CN in WES data. (A) Manhattan plot and quantile-quantile (QQ) plot for a gene-based rare variant association test of normalized, WES-measured MT-CN using WES data from both METSIM and FINRISK (N = 19,034). The red line represents a Bonferroni significance level of 2.164×10^−6^, as 23,105 genes were included in this test. *TMBIM1* is the only gene to reach significance at this level. (B) QQ plot for the test shown in (A). This plot is separated by minor allele frequency bin, as indicated by the colors and shapes of the points. (C) Boxplot showing the distributions of normalized WES-measured MT-CN, separated by the number of WES-detected alternate alleles in *TMBIM1* with MAF < 0.01 and CADD score > 20 (N = 19,034).

### Inference of causality in the association between MT-CN and insulin

To further understand the association between MT-CN and fasting serum insulin, we employed a Mendelian randomization (MR) approach with MT-CN as the exposure and insulin as the outcome. Using penalized regression, we leveraged our extensive phenotype data to build a genetic instrument from a large number of genetic variants and adjust for possible confounders via a novel approach (see Material and Methods; **Figure 4**). We believe this approach to be more robust to violations of key MR assumptions than other methods in situations where limited data are available and few robust genotype-exposure associations are known. We restricted our analysis to METSIM samples due to batch effects and inconsistencies in available quantitative trait data observed across FINRISK survey years (**Figure S4**). The effect sizes of the instrument in the causality test for insulin levels are shown in **Figure 4d**. We calculated our instrument using either L1 or L2 regularization. In both cases, the MT-CN instrument was not a significant predictor (α = 0.05) of insulin when we constructed our instrument from WGS variants, but was significant when the instrument was constructed from imputed array variants. This was likely due to the larger sample size of the imputed array data set. However, the effect estimates were remarkably similar across all four cases. As a result, inverse-variance weighted meta-analysis across datasets yielded highly significant P values for both penalties. In summary, our analysis provided evidence for a significant causal role for MT-CN in determining fasting serum insulin levels that was robust to the choice of regression penalty when building the genetic instrument. We note that this evidence for causality comes with some caveats (see Material and Methods).

### Replication and biological interpretation

In principle, changes in MT-CN can be caused by changes in the number of mitochondrial genome copies within cells or by changes in the blood cell type composition. Based on the association with rs9389268 and the nuances of the normalization procedure described above, we sought to test the hypothesis that our MT-CN measurement primarily reflects the cell type composition of the blood rather than the number of mitochondria per cell. We used imputed array genotype and phenotype data from the UK Biobank (N = 357,656) for this purpose^79^.

We first tested cell counts from the UK Biobank (UKBB) against a polygenic risk score (PRS) for MT-CN built using the genetic instrument from the Finnish data. Leukocyte, neutrophil, and platelet counts were all significantly associated with MT-CN PRS conditional on age, age^2^, and sex (see Material and Methods, **Table 6**). However, adjusting for neutrophil counts in the leukocyte regression eliminated the signal (PRS regression coefficient P = 0.839), suggesting that the leukocyte count signal was driven by the effect of neutrophil count. We removed any high leverage, large residual samples and repeated the neutrophil and platelet count regressions to ensure that this result was robust to outliers and found no appreciable change in significance (**Table 6**). As a result, we concluded that our MT-CN measurement was significantly associated with neutrophil and platelet counts. Subsequent analyses were performed both with and without adjustment for these variables, as described below.

**Table 6.**
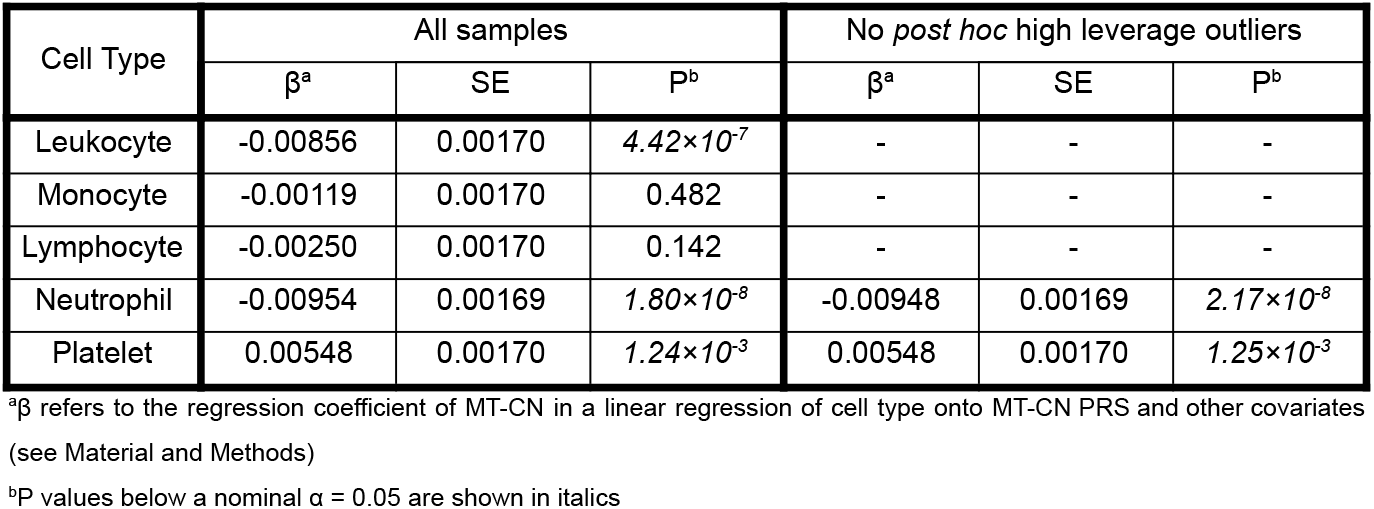
Association results between blood cell count traits and MT-CN polygenic risk score in 357,656 UK Biobank samples.

| Cell Type | All samples |  |  | No *post hoc* high leverage outliers |  |  |
| --- | --- | --- | --- | --- | --- | --- |
| | $\beta^a$ | SE | $P^b$ | $\beta^a$ | SE | $P^b$ |
| Leukocyte | -0.00856 | 0.00170 | *4.42×10<sup>-7</sup>* | - | - | - |
| Monocyte | -0.00119 | 0.00170 | 0.482 | - | - | - |
| Lymphocyte | -0.00250 | 0.00170 | 0.142 | - | - | - |
| Neutrophil | -0.00954 | 0.00169 | *1.80×10<sup>-8</sup>* | -0.00948 | 0.00169 | *2.17×10<sup>-8</sup>* |
| Platelet | 0.00548 | 0.00170 | *1.24×10<sup>-3</sup>* | 0.00548 | 0.00170 | *1.25×10<sup>-3</sup>* |

<sup>a</sup> $\beta$ refers to the regression coefficient of MT-CN in a linear regression of cell type onto MT-CN PRS and other covariates (see Material and Methods)
<sup>b</sup> $P$ values below a nominal $\alpha = 0.05$ are shown in italics

We note that the effect directions of the associations of platelet counts with metS and MT-CN PRS seem inconsistent at first glance, as platelet counts were positively correlated with MT-CN PRS (**Table 6**) and metS (**Table S3**) while MT-CN and insulin (a proxy for metS) were negatively correlated (**Figure 1b**). However, the FinMetSeq regression model in **Figure 1b** was not conditional on any other covariates (although age, age^2^, and sex were regressed out of the MT-CN measurement prior to this analysis), while the UKBB models that gave rise to **Table 6** and **Table S3** adjusted for many additional covariates, including 20 PCs and age-sex interaction terms. As a result, the effect directions for the analyses in the two datasets are not directly comparable.

We next tested for associations between MT-CN PRS and several cardiometabolic phenotypes from the test in **Figure 1a** (see Material and Methods). With the exception of C-reactive protein, which showed no significant association, all tested phenotypes showed nominal association with MT-CN PRS at α = 0.05, with total triglycerides and HDL being the only traits surviving Bonferroni correction (**Table 7**). We interpret this as replication of the link between mitochondrial genome copy number and metabolic syndrome in a large, independent data set.

**Table 7.** Association results between metabolic syndrome traits and MT-CN polygenic risk score in 357,656 UK Biobank samples.

| Trait | Without platelet and neutrophil adjustment |  |  | With platelet and neutrophil adjustment |  |  |
| --- | --- | --- | --- | --- | --- | --- |
| | $\beta^b$ | SE | $P^c$ | $\beta^b$ | SE | $P^c$ |
| Type 2 Diabetes | -0.1681 | 0.0826 | *0.0419* | -0.1467 | 0.0844 | 0.0823 |
| BMI | -0.0372 | 0.0175 | *0.0331* | -0.0233 | 0.0175 | 0.1836 |
| Fat Mass | -0.0452 | 0.0176 | *0.0100* | -0.0318 | 0.0177 | 0.0716 |
| C-Reactive Protein | -0.0015 | 0.0178 | 0.9305 | 0.0206 | 0.0171 | 0.2291 |
| HDL | 0.0573 | 0.0187 | *0.0021* | 0.0416 | 0.0187 | *0.0262* |
| Total Triglycerides | -0.0500 | 0.0176 | *0.0046* | -0.0330 | 0.0175 | 0.0603 |
| Weight<sup>a</sup> | -0.0359 | 0.0176 | *0.0414* | -0.0218 | 0.0178 | 0.2189 |

<sup>a</sup>The weight phenotype tested was that which was measured at the time of impedance measurement.
<sup>b</sup> $\beta$ refers to the regression coefficient of MT-CN in a linear regression of cell type onto MT-CN PRS and other covariates (see Material and Methods)
<sup>c</sup> $P$ values below a nominal $\alpha = 0.05$ are shown in italics

To determine whether there was any association between MT-CN and metabolic syndrome not mediated through cell counts, we repeated the tests of cardiometabolic trait association with MT-CN PRS with adjustment for platelet and neutrophil counts. HDL was the only trait with a nominal (α = 0.05) association with PRS under this adjustment, but this signal was not strong enough to survive Bonferroni correction (**Table 7**). This suggests that the associations we observed between MT-CN and metabolic traits arose simply because MT-CN is a proxy for platelet and neutrophil count. This was supported by the fact that direct testing of platelets and neutrophils against triglycerides, fat mass, and HDL yielded remarkably significant associations, which survived *post-hoc* removal of high-leverage, high-residual outlier samples (**Table S3**). This evidence for MT-CN as a proxy for platelet and neutrophil counts strongly suggests that the causal relationship observed in the Mendelian randomization experiment (see above) in fact represents a causative role for neutrophils and platelet counts in setting serum insulin levels.

Given the strong observed associations between blood cell count phenotypes and MT-CN PRS, we used these blood phenotypes to seek replication of the genetic associations detected in Finnish data. Using imputed UKBB genotype data, we tested the expected alternate allele dosage of both rs2288464 and rs9389268 against the same blood cell traits mentioned above, using linear regression (**Table S4**; expected alternate allele dosage was calculated from genotype call probabilities as *DS* = *P* (0*/*1) + 2*P* (1*/*1)). As expected given its known associations with multiple hematological parameters (see above), rs9389268 showed strong associations with all tested blood cell phenotypes. rs2288464 was not significantly associated with any of the five phenotypes after correction for multiple testing, although a nominal association was detected with total leukocyte count. This further strengthens our belief that rs9389268 is truly associated with MT-CN through blood cell composition. We also tested *TMBIM1* against the same blood cell traits in UKBB using SKAT-O^48^, and found no significant associations (**Table S4**). This may mean that *TMBIM1* affects MT-CN through a mechanism other than altering blood cell type composition.

As further evidence that MT-CN is a proxy for blood cell composition, we looked up MT-CN association P values in METSIM for the top five neutrophil and platelet count QTLs from the NHGRI-EBI GWAS Catalog. Out of ten variants tested, five had P < 0.05 in METSIM (**Table S11**). We note that three of these five were either near or identical to known MT-CN loci (including rs9389268, the marker identified in this study). rs25645, a variant reported to be highly associated with neutrophil count^68^, is only 2.5 kb away from rs709591, a SNP with a reported suggestive association with MT-CN^27^ and a P value of 1.61×10^−4^ in our METSIM study. Moreover, rs11759553, a platelet-associated variant^68^, is 324 kb away from rs9389268, the lead marker for MT-CN in METSIM (rs11759553 P = 2.15×10^−10^ in METSIM). Finally, rs445 was reported as a lead marker for both MT-CN association^25^ and platelet count^68^. rs445 has P = 0.048 for association with MT-CN in METSIM. While none of the 10 known cell count-associated markers tested achieved significance beyond a Bonferroni threshold, the overlap between these variants and independently-measured MT-CN QTLs was suggestive of a relationship between cell counts and whole blood-derived MT-CN.

Using UKBB data, we further sought to generate hypotheses for other phenotypic associations with MT-CN. To this end, we performed a phenome-wide screen of MT-CN PRS against all of the UKBB phenotypes available to us. To curate and transform these phenotypes, we used a modified version of PHESANT^61,62^, which outputs all continuous variables in both raw and inverse rank-normalized form. We chose to interpret the results from the normalized continuous variables (**Table S5**) to be conservative and robust to outliers, although the results of the raw continuous variable analyses were similar (**Table S6**). No metabolic syndrome traits appeared among the tested traits with q < 0.05. However, the tests for HDL cholesterol, self-reported heart attack, and doctor-diagnosed heart attack did yield somewhat suggestive results (q = 0.123, 0.176, and 0.176, respectively). We also repeated this screen with adjustment for neutrophil and platelet counts (**Table S7** and **Table S8**), resulting again in no metabolic syndrome phenotypes achieving *q* < 0.05. The addition of neutrophil and platelet counts as covariates attenuated the suggestive signals for HDL cholesterol, self-reported heart attack, and doctor-diagnosed heart attack (q = 0.284, 0.391, and 0.402, respectively).

## Discussion

We have described one of the most well-powered studies to date of the genetic relationship between MT-CN measurements in blood and cardiometabolic phenotypes. Our study is one of very few of which we are aware to utilize WGS data, found to be the most reliable method for estimating MT-CN in a recent study^18^, for this purpose. Our data show highly significant associations between blood-derived MT-CN measurements and several cardiometabolic traits, particularly insulin and fat mass. We observed strong heritability of MT-CN (31%), on par with other widely studied cardiometabolic traits such as LDL, and identified one single marker association on a haplotype previously associated with several hematological parameters^71–74^. A previous study using qPCR to quantify MT-CN reported two sub-threshold QTLs^27^; of these markers, only rs709591 replicated in our study (P = 1.61×10^−4^). We also report one gene with a rare-variant association with MT-CN, *TMBIM1*, that has a known link to non-alcoholic fatty liver disease^78^. More work is needed to replicate this genetic association.

Using a novel multiple-variant instrument-building method, we report evidence from Mendelian randomization supporting a causal role for MT-CN in metabolic syndrome. Further, we used UK Biobank data to show that not only does the link between MT-CN and metabolic syndrome replicate in an independent data set using a polygenic risk score approach. Contrary to previous claims that variability in the number of mitochondria per cell is responsible for CHD risk^12^, this association is mediated by neutrophil and platelet counts.

One important question that our study cannot definitively resolve is the relative contribution of intracellular mitochondrial abundance versus cell-type composition differences in determining the measured MT-CN value. We identified a MT-CN association result at a known QTL for cell type composition of blood^71–74^ (HBS1L-MYB), and we further replicated a prior sub-threshold association at a different neutrophil-associated locus^67,68^ (rs709591). Together, these results argue that cell type composition is an important component of this measurement. On the other hand, two other significant associations from the Finnish dataset (rs2288464, *TMBIM1*) showed no effect on cell type composition in the UK Biobank. Future work in large cohorts with both WGS and cell count data – which were not simultaneously measured in any samples in this study – will be required to rigorously determine what blood-derived MT-CN primarily measures. However, the results of our MR and UK Biobank analyses together suggest that MT-CN is causally related to metabolic syndrome traits, and that this relationship is mediated by cell-type composition differences.

There is prior evidence to support the role of inflammation – specifically via innate immune cells such as neutrophils – in the etiology of type 2 diabetes (T2D) and insulin resistance^80–82^, which suggests a plausible model by which peripheral blood neutrophil count could influence metabolic syndrome. Nutrient excess and high fat diets are known to recruit neutrophils into tissues, which then cause insulin resistance both by releasing TNF-α and IL-6 and by upregulating cyclooxygenase^80^. This leads to increased LTB4 and subsequent upregulation of NF-κB, a central regulator of inflammation. Moreover, free fatty acids also cause neutrophils to stay in tissues longer, resulting in persistent inflammation and leading to insulin resistance^80^. While it is known that inflammation, and particularly neutrophils, play a role in metabolic syndrome, our results strongly suggest that peripheral blood neutrophil count causally contributes to this process and is associated with heritable genetic variation in the human population. Overall, our work provides further insight into the role that inflammation plays in metabolic syndrome and supports the idea that targeting inflammation may be a fruitful avenue of investigation in developing future therapeutics.

## Supporting information

Large Supplemental Tables

## Data Availability

METSIM WGS, METSIM WES, and FINRISK WES sequence data are available through dbGaP (accessions phs001579, phs000752, and phs000756). METSIM variant data as well as MT-CN phenotype values will soon be available through AnVIL
(accessions TBD). Imputed array GWAS summary statistics from METSIM and WES SKAT-O summary statistics from the joint dataset are freely available at https://wustl.box.com/s/7xfbmxq2r4kg8p8bfc7vpqlrmqvhm0lx. Genomic and phenotypic data for the FINRISK cohort are or will soon be obtainable through THL Biobank, the Finnish Institute for Health and Welfare, Finland (https://thl.fi/en/web/thl-biobank).

## Supplemental Information Description

Supplemental data include 8 figures and 11 tables.

## Acknowledgements

This work was funded by the NHGRI Centers for Common Disease Genetics (grant number UM1 HG008853 to I.M.H. and N.O.S), the NHGRI large-scale sequencing grant (grant number 5U54HG003079), the Sigrid Jusélius Foundation (to S.R.), the University of Helsinki HiLIFE Fellow grants 2017-2020 (to S.R.), the Academy of Finland Center of Excellence in Complex Disease Genetics (grant number 312062 to S.R.), the Academy of Finland (grant number 285380 to S.R.), the National Heart, Lung and Blood Institute (grant number T32HL007081 to E.Y.), and the National Center for Advancing Translational Sciences (grant number UL1TR002345 to E.Y.).

We thank Hyeim Jung for her help in identifying outlier individuals as well as the WashU data production team, in particular Robert Fulton, Lucinda Fulton, Catrina Fronick, Aye Wollam and Susan K. Dutcher. This research has been conducted using the UK Biobank Resource under Application Number 56546. The FINRISK samples used for the research were obtained from the FINRISK Study and from THL Biobank. We thank all study participants for their generous participation at THL Biobank and the FINRISK Study.

## Declaration of Interests

N.O.S has received grant funding from Regeneron Pharmaceuticals for unrelated work. The rest of the authors declare no competing interests.

### Web Resources

EPACTS, https://genome.sph.umich.edu/wiki/EPACTS

lmPerm, https://github.com/mtorchiano/lmPerm

NHGRI-EBI GWAS Catalog, https://www.ebi.ac.uk/gwas

smartPCA, https://github.com/DReichLab/EIG

## Data and Code Availability

METSIM WGS, METSIM WES, and FINRISK WES sequence data are available through dbGaP (accession numbers phs001579, phs000752, and phs000756). METSIM callsets from WGS and imputed array data as well as MT-CN phenotype values will soon be available through AnVIL. Imputed array GWAS summary statistics from METSIM and and WES SKAT-O summary statistics from the joint dataset are freely available at https://wustl.box.com/s/7xfbmxq2r4kg8p8bfc7vpqlrmqvhm0lx. Genomic and phenotypic data for the FINRISK cohort are obtainable through THL Biobank, the Finnish Institute for Health and Welfare, Finland (https://thl.fi/en/web/thl-biobank).

