## Supplemental Information for "Mitochondrial genome copy number measured by DNA sequencing in human blood is strongly associated with metabolic traits via cell-type composition differences"

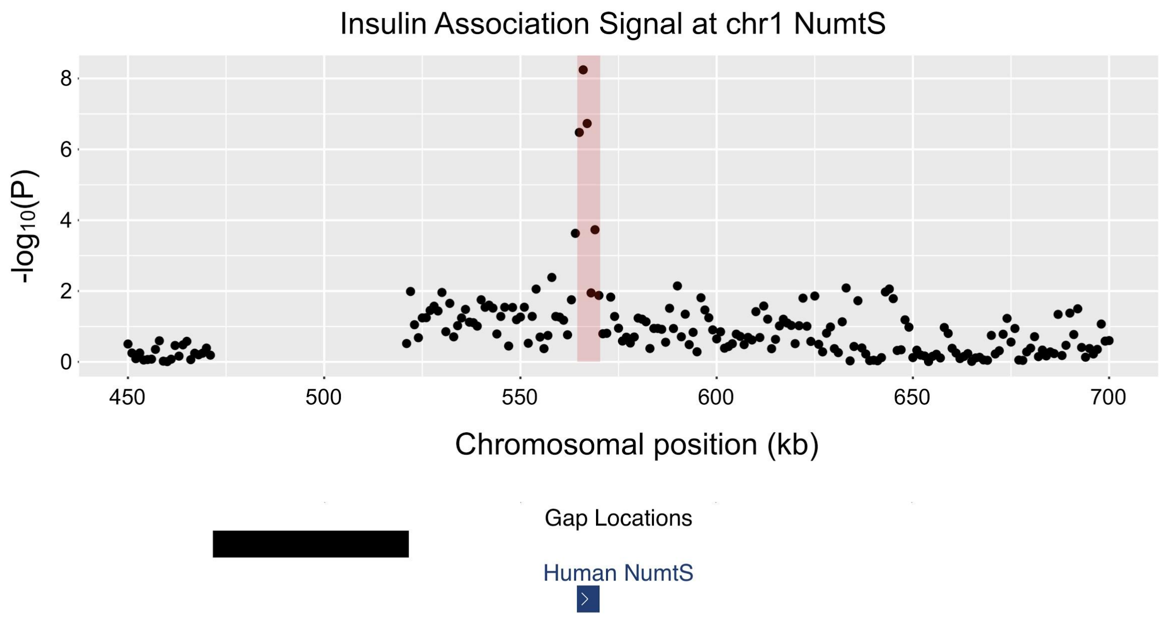


**Figure S1. Insulin association signal at chromosome 1 NumtS.** Manhattan plot from an analysis of copy number variation at nuclear mitochondrial insertion sites (NumtS) using CNVnator read-depth measurements in 1 kb genomic windows, using an initial set of 2,049 samples from an earlier data freeze analyzed for an unrelated CNV association study (manuscript under review - preprint doi: 10.1101/2020.12.13.422502). Shown at the bottom are two tracks from the UCSC Genome Browser. The black track represents an assembly gap upstream of the association signal, and the blue track shows the location of the NumtS region where the association peak is located.

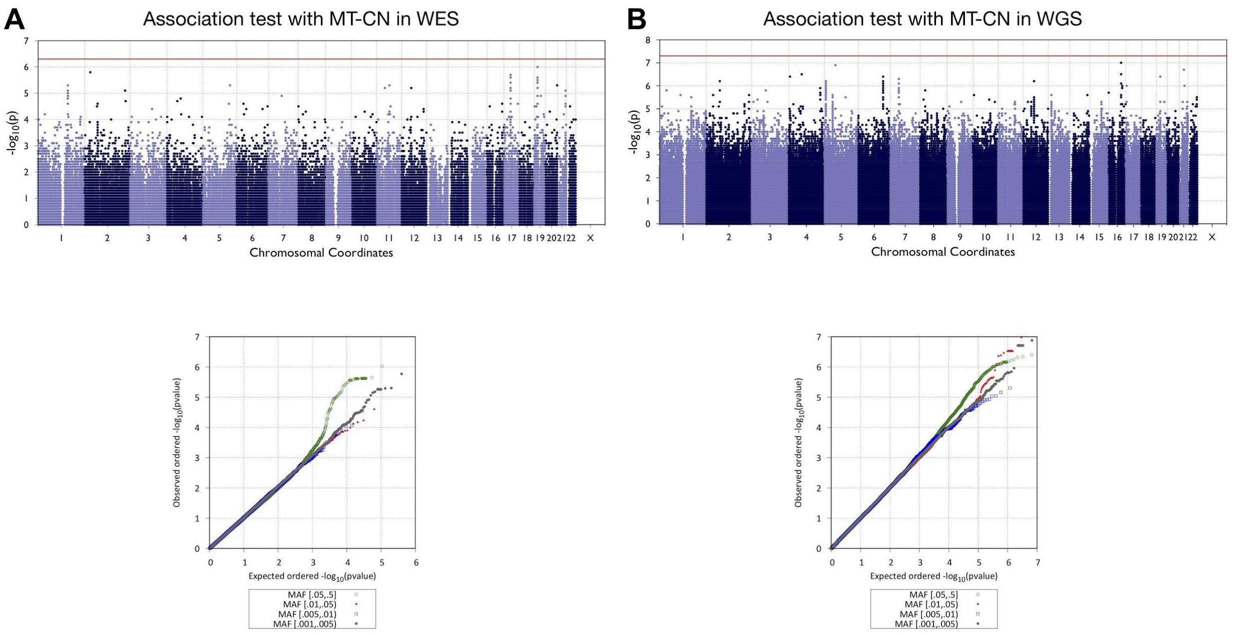


**Figure S2. Joint (METSIM and FINRISK) single-marker association tests with MT-CN in WGS and WES data.** (A) Manhattan plot and quantile-quantile (QQ) plot for an exome-wide association test of normalized, WES-measured MT-CN using WES genotype data (N = 19,034). The red line represents an exome-wide significance level of $5{\times}10^{-7}$; no tested markers achieved this level of significance. The QQ plot is separated by minor allele frequency bin, as indicated by the colors and shapes of the points. (B) Manhattan plot and quantile-quantile (QQ) plot for a genome-wide association test of normalized, WGS-measured MT-CN using WGS genotype data (N = 4,149). The red line represents a genome-wide significance level of $5{\times}10^{-8}$; no tested markers achieved this level of significance. The QQ plot is separated by minor allele frequency bin, as indicated by the colors and shapes of the points.

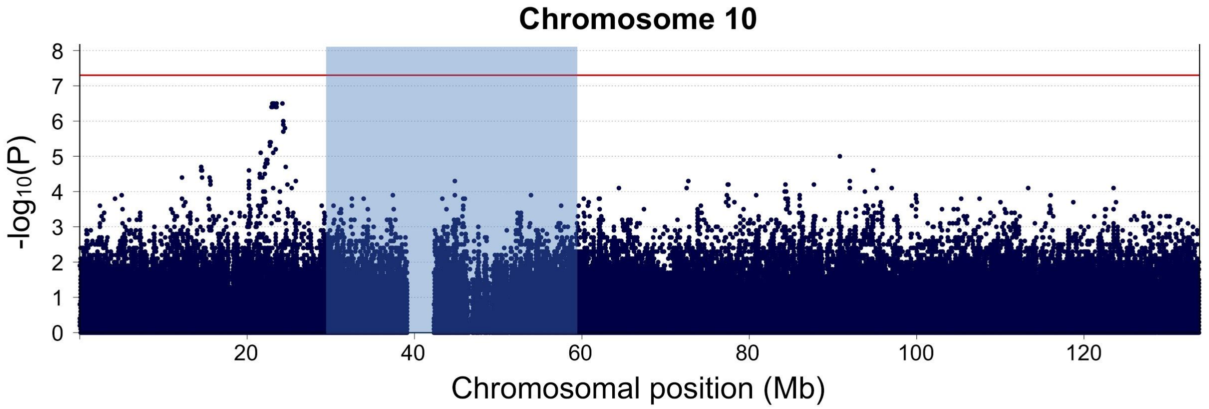


**Figure S3. Single-marker association test results at chromosome 10 QTL identified by Curran *et al.*** Chromosome 10 Manhattan plot for association with WES-estimated MT-CN in Finnish imputed array data from METSIM (N = 9,791). The blue highlighted region represents the approximate location of the linkage peak reported for MT-CN in Curran *et al.*, 2007. This region is larger than the reported region in the Curran study (~30 Mb vs. ~24 Mb) because the exact coordinates of the 1-LOD support interval were not reported explicitly, so the genetic coordinates were approximated from a figure and converted to physical coordinates using a genetic map from HapMap Phase 2. Some error may have been introduced due to differing populations between the HapMap data and the Mexican-American samples used by Curran.

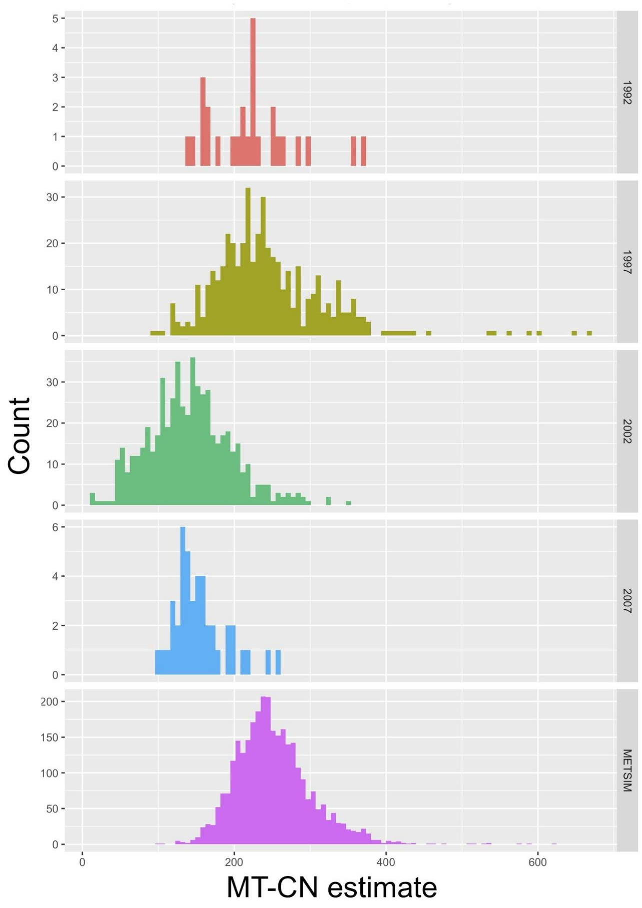


**Figure S4. Raw WGS-based MT-CN estimate distributions for METSIM and all four FINRISK surveys.** Facet labels "1992", "1997", "2002", and "2007" refer to individual FINRISK survey years.

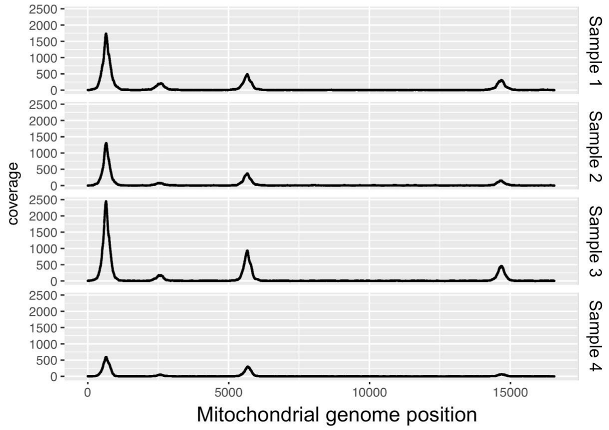


**Figure S5. Nonuniform WES coverage across the mitochondrial genome for four representative samples.**

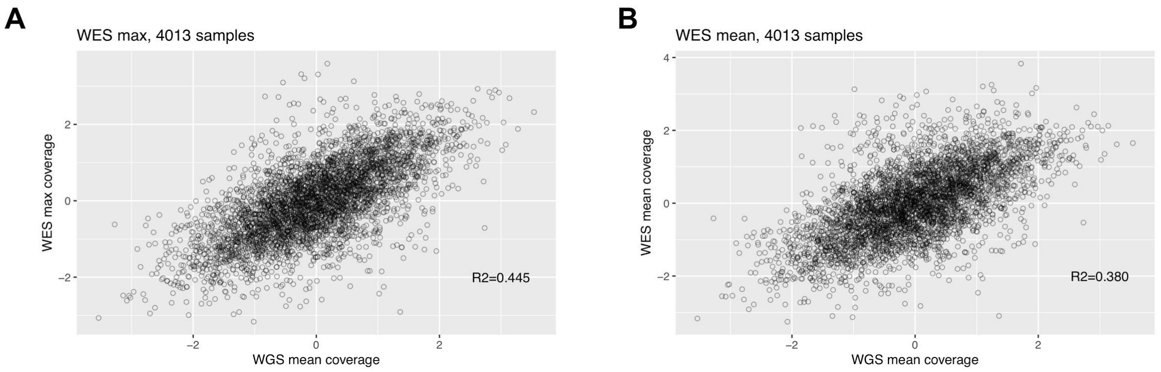


**Figure S6. Comparison of two methods of summarizing nonuniform mitochondrial coverage in WES into a single measurement.** Both panels show the correlation with WGS mean coverage for the 4,013 samples for which both WGS and WES data were available. (A) shows the results of summarizing WES mitochondrial coverage using the maximum coverage value across the mitochondrial chromosome while (B) shows the results of using the mean mitochondrial coverage instead. $R^2$ values are shown on each panel.

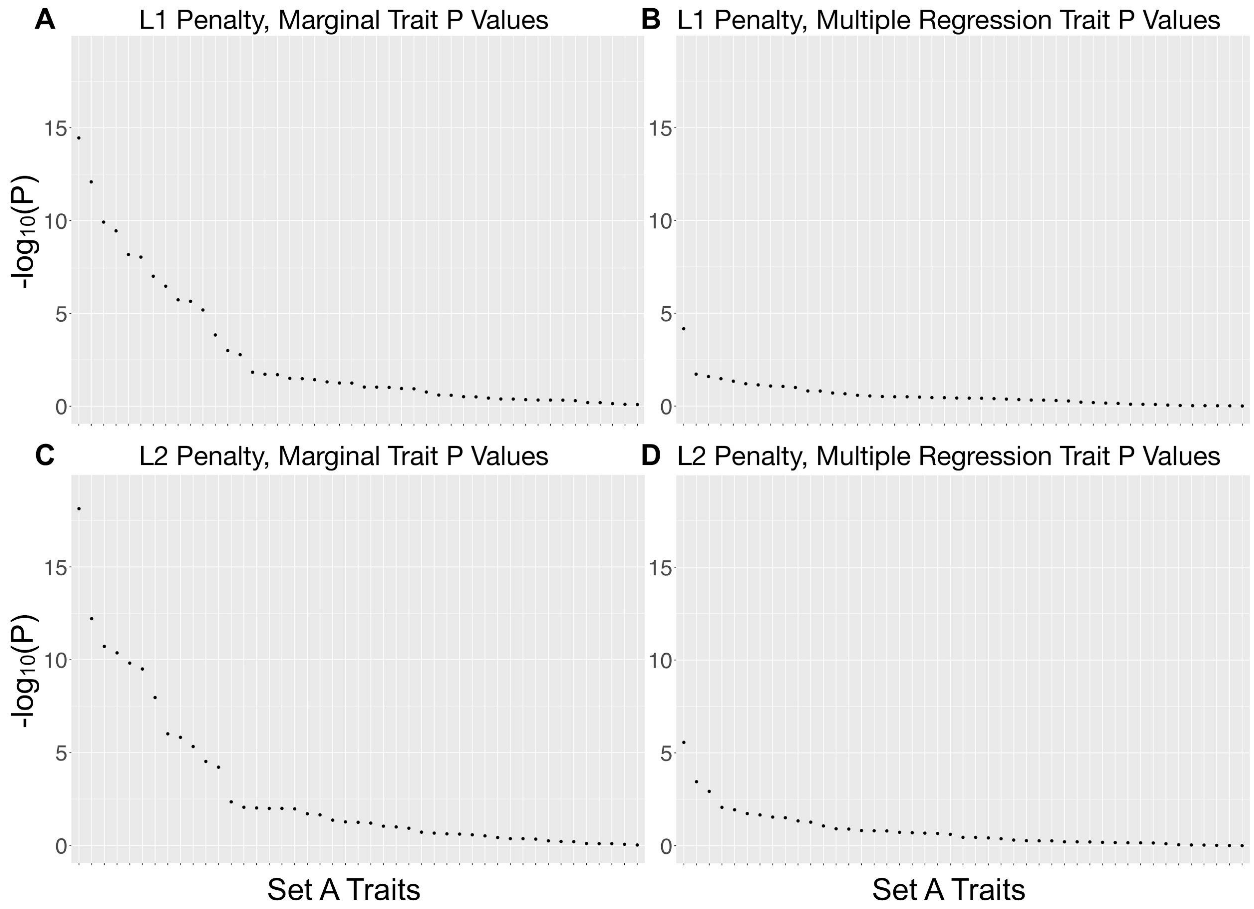


**Figure S7. Associations between Z and traits in set A in imputed data**. P values computed from linear regression using the output from a randomly chosen run of multiple imputation of phenotype data. (A) and (C) show the marginal P values of the traits in set $A$ from regressions of $Z$ onto each trait separately, while (B) and (D) show the P values from multiple regression of $Z$ onto all traits in set $A$. The instrument in (A) and (B) was computed using an L1 penalty, while that in (C) and (D) was computed using an L2 penalty.

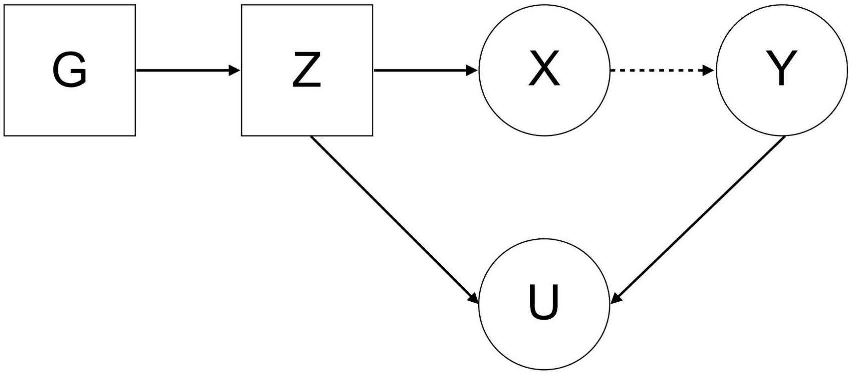


**Figure S8. Example of collider bias.** If Z and Y are independently causal for a variable U, then adjusting for U can induce an association between Z and Y that did not previously exist.

| | All WES samples | | | | Samples with WGS and WES data | | | | WES samples excluding WGS samples | | | |
|---|---|---|---|---|---|---|---|---|---|---|---|---|
| | N | Beta | SE | P | N | Beta | SE | P | N | Beta | SE | P |
| Fat mass | 11577 | -0.094 | 0.010 | 5.79E-23 | 3016 | -0.102 | 0.017 | 2.25E-09 | 8561 | -0.091 | 0.011 | 3.01E-15 |
| ln(Fasting insulin) | 9434 | -0.127 | 0.010 | 1.10E-33 | 2689 | -0.135 | 0.019 | 1.98E-12 | 6745 | -0.124 | 0.013 | 1.29E-22 |

**Table S2. MT-CN associations with insulin and fat mass in WES data.** Results of EMMAX test of association between normalized MT-CN and both fat mass and fasting serum insulin using WES data. Association tests were performed in all samples and also separately among samples with and without WGS data.

| | Fat Mass | | | HDL | | | Total Triglycerides | | |
|---|---|---|---|---|---|---|---|---|---|
| | Beta | SE | P | Beta | SE | P | Beta | SE | P |
| Platelet count | 0.044 | 0.002 | 6.01E-144 | -0.019 | 0.002 | 1.36E-26 | 0.088 | 0.002 | <2.22E-308 |
| Neutrophil count | 0.139 | 0.002 | <2.22E-308 | -0.142 | 0.002 | <2.22E-308 | 0.179 | 0.002 | <2.22E-308 |
| Platelet count (no *post-hoc* outliers) | 0.044 | 0.002 | 5.58E-144 | -0.019 | 0.002 | 1.19E-26 | 0.088 | 0.002 | <2.22E-308 |
| Neutrophil count (no *post-hoc* outliers) | 0.139 | 0.002 | <2.22E-308 | -0.142 | 0.002 | <2.22E-308 | 0.179 | 0.002 | <2.22E-308 |

**Table S3. Direct associations between platelet/neutrophil counts and metabolic syndrome phenotypes in the UK Biobank.** Results of direct testing of platelet and neutrophil counts against metabolic syndrome phenotypes chosen based on marginally significant associations for MT-CN without adjustment for cell counts. Testing was done in the UK Biobank (N = 357,656) using linear regression of traits onto cell counts conditional on the same covariates as the MT-CN analyses (see Material and Methods). The bottom two rows show the results after removal of high-residual, high-leverage outliers as determined by Cook’s distance.

| | rs2288464 | | | rs9389268 | | | *TMBIM1* |
|---|---|---|---|---|---|---|---|
| | Beta | SE | P | Beta | SE | P | P |
| Leukocyte | 0.00921 | 0.00406 | 0.023 | -0.0471 | 0.0027 | 4.85E-66 | 0.784 |
| Monocyte | 0.00670 | 0.00406 | 0.099 | -0.0331 | 0.0027 | 2.18E-33 | 0.331 |
| Lymphocyte | 0.00525 | 0.00407 | 0.197 | -0.0372 | 0.0028 | 1.65E-41 | 1 |
| Neutrophil | 0.00703 | 0.00405 | 0.083 | -0.0363 | 0.0027 | 5.66E-40 | 0.274 |
| Platelet | 0.00714 | 0.00406 | 0.079 | 0.1109 | 0.0027 | <2.22E-308 | 0.082 |

**Table S4. Associations in the UK Biobank between cell counts and loci mapped to MT-CN in Finnish data.** Association tests in the UK Biobank (N = 357,656) of cell counts against common-variant and gene-based rare-variant associations found in Finnish GWAS. Single marker association statistics are calculated by linear regression, while *TMBIM1* P values are calculated by SKAT-O.

| Dataset | n | Penalty | Beta | Std. Error | P |
|---|---|---|---|---|---|
| WGS | 3034 | L1 | -0.019 | 0.013 | 0.1471 |
| | | L2 | -0.020 | 0.011 | 0.0822 |
| Imputed Array | 6774 | L1 | -0.019 | 0.008 | 0.0188 |
| | | L2 | -0.023 | 0.008 | 0.0045 |
| Meta-analysis | - | L1 | -0.019 | 0.007 | 0.0057 |
| | | L2 | -0.022 | 0.007 | 0.0009 |

**Table S9. Mendelian randomization results after removal of high leverage *post hoc* outliers.** Results of Mendelian randomization test for causality of MT-CN on fasting serum insulin, after removal of high leverage *post hoc* outliers (see “Inference of causality in the association between MT-CN and insulin”). These results are nearly identical to those computed before outlier removal (**Figure 4d**), indicating that the detected signals were not driven by outlier samples.

| Trait | SNP | GRCh37 coordinates | Ref. | Nearest gene(s) | METSIM MAF | METSIM P value | Notes |
|---|---|---|---|---|---|---|---|
| MT-CN | rs445 | 7:92408370 | Cai, *et al.*[1] | CDK6 | 0.0504 | 0.04842 | Lead marker for MT-CN in Cai, *et. al*[1] and neutrophil count in Chen *et. al*[2] |
| | rs11006126 | 10:60142880 | | TFAM | 0.100 | 0.3609 | |
| | rs709591 | 17:38175561 | Guyatt *et al.*[3] | CSF3, MED24, PSMD3 | 0.402 | 1.61E-04 | Not genome-wide significant in literature; MED24 is associated with neutrophil count (Chen et al.[2]) |
| | rs12873707 | 13:104810437 | | LOC105370340 | 0.061 | 0.5048 | Not genome-wide significant in literature |
| Neutrophil count | rs25645 | 17:38173143 | Chen *et al.*[2] | CSF3 | 0.322 | 0.01712 | 2.5 kb away from rs709591 |
| | rs2814778 | 1:159174683 | | ACKR1, CADM3-AS1 | 0.00056 | 0.6244 | |
| | rs9131 | 4:74963049 | | CXCL2 | 0.355 | 0.04622 | |
| | rs56388170 | 7:28724374 | | CREB5 | 0.299 | 0.009071 | |
| | rs445 | 7:92408370 | | CDK6 | 0.0504 | 0.04842 | Lead marker for MT-CN in Cai, *et. al*[1] and neutrophil count in Chen *et. al*[2] |
| Platelet count | rs11759553 | 6:135422296 | Chen *et al.*[2] | HBS1L | 0.348 | 2.15E-10 | 324 kb away from rs9389268, lead marker for MT-CN in METSIM |
| | rs9861033 | 3:56861222 | | ARHGEF3 | 0.339 | 0.2397 | |
| | rs113608931 | 9:4758972 | | AL353151.2 | 0.301 | 0.3665 | |
| | rs11066309 | 12:112883476 | | PTPN11 | 0.379 | 0.1081 | |
| | rs549888 | 6:33552202 | | GGNBP1 | 0.453 | 0.1007 | |

**Table S11. Replication in METSIM of published MT-CN QTLs**. Results of GWAS of METSIM using imputed genotypes (N = 9,791) for top hits in the literature for MT-CN, neutrophil count, and platelet count. MT-CN loci were taken from references Cai, *et al*.[1] and Guyatt *et al.*[3]. Neutrophil and platelet count loci were chosen as the top 5 loci in the NHGRI-EBI GWAS Catalog for their respective traits, with some curation to ensure the loci tested were mutually independent. All ten variants came from either trans-ethnic or European-specific GWAS reported in reference Chen *et al*.[2]
